# Multimodal Analysis of Secondary Cerebellar Alterations after Pediatric Traumatic Brain Injury

**DOI:** 10.1101/2022.12.24.22283926

**Authors:** Finian Keleher, HM Lindsey, Rebecca Kerestes, Houshang Amiri, Robert F Asarnow, Talin Babikian, Brenda Bartnik-Olson, Erin D Bigler, Karen Caeyenberghs, Carrie Esopenko, Linda Ewing-Cobbs, Christopher C Giza, Naomi J Goodrich-Hunsaker, Cooper B Hodges, Kristen R Hoskinson, Andrei Irimia, Marsh Königs, Jeffrey E Max, Mary R Newsome, Alexander Olsen, Nicholas P Ryan, Adam T Schmidt, Dan J Stein, Stacy J Suskauer, Ashley L Ware, Anne Wheeler, Brandon A Zielinski, Paul M Thompson, Ian Harding, David F Tate, Elisabeth A Wilde, Emily L Dennis

**Author notes:** Correspondence to: Dr. Emily L Dennis, TBI and Concussion Center, Dept of Neurology, University of Utah School of Medicine.

## Abstract

While traditionally ignored as a region purely responsible for motor function, the cerebellum is increasingly being appreciated for its contributions to higher order functions through various cerebro-cerebellar networks. Traumatic brain injury (TBI) research generally focuses on the cerebrum, in part because acute pathology is not found in the cerebellum as often. Acute pathology is an important predictor of outcome, but neural disruption also evolves over time in ways that have implications for daily-life functioning. Here we examine these changes in a multi-modal, multi-cohort study.

Combining 12 datasets from the Enhancing NeuroImaging Genetics through Meta-Analysis (ENIGMA) Pediatric msTBI (moderate-severe TBI) working group, we measured volume of the total cerebellum and 17 subregions using a state-of-the-art, deep learning-based approach for automated parcellation in 598 children and adolescents with or without TBI (msTBI; n = 314 | non-TBI; n = 284; age M = 14.0 ± 3.1 years). Further, we investigated brain-behavior relations between cerebellar volumes and a measure of executive functioning (i.e., Behavioral Rating Inventory of Executive Function [BRIEF]). In a subsample with longitudinal data, we then assessed whether late changes in cerebellar volume were associated with early white matter microstructural organization using diffusion tensor imaging (DTI).

Significantly smaller total cerebellar volume was observed in the msTBI group (Cohen’s *d* = −0.37). In addition, lower regional cerebellar volume was found in posterior lobe regions including crus II, lobule VIIB, lobule VIIIB, vermis VII, and IX (Cohen’s *d* range = −0.22 to −0.43). Smaller cerebellum volumes were associated with more parent-reported executive function problems. These alterations were primarily driven by participants in the chronic phase of injury (> 6 months). In a subset of participants with longitudinal data (n = 80), we found evidence of altered growth in total cerebellum volume, with younger msTBI participants showing secondary degeneration in the form of volume reductions, and older participants showing disrupted development reflected in slower growth rates. Changes in total cerebellum volume over time were also associated with white matter microstructural organization in the first weeks and months post-injury, such that poorer white matter organization in the first months post-injury was associated with decreases in volume longitudinally.

Pediatric msTBI was characterized by smaller cerebellar volumes, primarily in the posterior lobe and vermis. The course of these alterations, along with group differences in longitudinal volume changes as well as injury-specific associations between DTI measures and volume changes, is suggestive of secondary cerebellar atrophy, possibly related to supra-tentorial lesions, and/or disruption in cerebellar structural and functional circuits. Moreover, evidence for robust brain-behavior relationships underscore the potential cognitive and behavioral consequences of cerebellar disruption during a critical period of brain development.

## Introduction

Children and adolescents are particularly vulnerable to the consequences of traumatic brain injury (TBI). In recent years, TBI has been a leading cause of death and disability in children ages 1-18 years old in the United States, with annual associated medical costs in excess of $1 billion (CDC, 2021).^1^ Pediatric TBI, as compared to adult TBI, is associated with several distinct characteristics due to age-related developmental, anatomical, and physiological differences.^2^ Researchers have posited two contrasting working models to explain these findings: either 1) that the immature brain is more plastic, and thus more flexible and able to adapt following insult, and/or 2) that the immature brain is uniquely vulnerable, and thus more susceptible to developmental disruption.^3^ Both models emphasize the significance of varying maturational trajectories in determining outcome. Most studies of pediatric TBI have therefore targeted supratentorial brain areas that are thought to be later-developing and/or more vulnerable to direct injury. However, recent research suggests that both of these models may be inaccurate or incomplete.

To date, there has been a paucity of pediatric TBI research on the cerebellum because of its purported early maturation and relatively lower mechanical vulnerability.^4^ However, novel image processing tools and more fine-grained atlases have allowed researchers to segment the cerebellum into subregions, or lobules, and measure regional volumes over the lifespan.^5^ These studies have found complex developmental trajectories for cerebellar subregions, with maturation trending in the anterior to posterior direction. Vermal regions reach peak maturation before age 5 years,^6^ anterior lobe (lobules I-V) between ages 12-16, while the maturational peaks for the posterior lobe (lobules VI-IX) span adolescence and early adulthood (other than lobule IX, which matures around age 5 years).^7^ As such, many subregions of the cerebellum are in critical periods of development during adolescence, which may make them especially vulnerable to developmental disruption as a consequence of pediatric TBI. Additionally, the cerebellum’s role in higher order cognitive abilities is driven by connectivity with cerebral regions such as the prefrontal cortex. As these cerebral regions develop, so too do their connections with the cerebellum.

Traditionally, neuroimaging studies of TBI have focused on supratentorial regions and white matter tracts due to their vulnerability to traumatic/diffuse axonal injury (TAI/DAI). Diffusion tensor imaging (DTI) can detect differences in myelination and axon density, and is sensitive to TAI. In recent years, alterations have been observed in the cerebellum, including decreases in white matter volume,^8^ reductions in fractional anisotropy (FA),^9^ functional dissociation,^10^ and hypoperfusion.^11^ The mechanisms underlying these alterations remain unclear, and while such effects are often due to direct injury, such injury is less likely in the cerebellum because of its location in the brain.^12^ An alternative source of cerebellar alteration may be “connectomal diaschisis,”^13^ whereby direct injury to the cerebrum may propagate to the cerebellum via cerebellar structural and functional networks.^14,15^ Animal research supports this possibility, with studies showing indirect alterations to the cerebellum related to disruptions in the fibers of cortico-cerebellar circuits.^16–18^ Assuming that this is the process through which most post-injury cerebellum disruption occurs, structural alterations would not be present in the first few months post-injury but rather would take months to develop.

The functional consequences of cerebellar injury are not fully understood. The cerebellum has long been associated with aspects of motor functioning, such as balance, coordination, motor learning, and body awareness,^19^ but fronto-cerebellar brain systems may also be involved in executive functions^20^ such as multitasking,^21^ inhibition,^22^ working memory,^23^ social cognition,^24^ and emotional processing.^25^ While several investigations of cerebellar injury in adult patients with TBI,^26^ brain tumor,^27^ or stroke ^28^ have supported these relationships, there are inconsistencies in the literature as some studies have shown no significant relationship between cerebellar morphological measures and executive functioning.^29,30^ These inconsistencies are likely due to small sample sizes, variability in the mechanism of injury, heterogeneity in neuropsychiatric measures, or developmental incongruity. Nevertheless, the possibility that injury-associated alterations to the cerebellum contribute to executive dysfunction demands further research, particularly following pediatric TBI, as no prior studies have examined the association between executive dysfunction and cerebellar morphological changes after pediatric TBI. More importantly, understanding mechanisms whereby injury can affect the cerebellum may be key in improving diagnosis, treatment, and outcomes in pediatric TBI.

The current study investigated volumetric cross-sectional differences and longitudinal changes in the cerebellum following pediatric mild complicated-severe TBI, and aimed to determine if these changes are associated with white matter microstructural organization and executive functioning. Enhancing Neuroimaging Genetics through Meta-Analysis (ENIGMA) is a global consortium of researchers aimed at achieving greater statistical power through harmonized processing of legacy data. Combining 12 cohorts shared within the ENIGMA Pediatric Moderate/Severe TBI (msTBI) working group,^31,32^ we measured regional cerebellar volume in a cohort of 598 children/adolescents, including those with mild complicated-severe TBI (msTBI; n = 314) or without TBI (non-TBI; n = 284) and examined its associations with white matter microstructure and executive functioning. We hypothesized that (1) cerebellar volume would be lower in participants with msTBI than the non-TBI group, that (2) these disruptions would be most prominent in patients furthest from the time of injury, and that (3) smaller cerebellar volume would be associated with poorer executive functioning.

## Materials and Methods

### Study Design

The ENIGMA Pediatric msTBI Working Group is a subgroup of the ENIGMA Brain Injury Working Group,^33,34^ a collaborative group of neuroimaging researchers around the world whose focus is TBI.^32^ ENIGMA works through collaborative, harmonized data processing and analysis, bringing together data from different sources to identify reliable neuroimaging biomarkers of injury and recovery. Our initial hypotheses focused on cerebellar volumes, but results from these analyses motivated us to include available DTI data, hypothesizing that alterations in DTI would predate and predict changes in cerebellar volumes.

### Study Samples

Our analysis consisted of 12 previously collected cohorts across 9 research sites, totaling 314 participants with TBI [ranging between complicated mild-severe TBI (msTBI); complicated mild = mild (Glasgow Coma Scale [GCS]>12) but with injury-related imaging abnormalities] and 284 non-TBI participants (demographic details in **Table 1**). The non-TBI group consisted of healthy (HC; n = 133) and orthopedically injured (OI; n = 151) children. Participants with msTBI or OI were recruited from hospitals and outpatient rehabilitation facilities, while healthy controls were recruited from the surrounding communities. Further details on recruitment and imaging parameters for each site may be found in **Supplementary Tables 1** and **2**, respectively. Across cohorts, the age range of the sample was 5.5-19.7 years (M = 14.0 ± 3.1). In line with prior publications,^31^ we divided msTBI participants into three post-injury windows: (i) acute/sub-acute (MRI within 7 weeks of injury), during which pathology such as intracerebral hemorrhage and edema dominate; (ii) post-acute (MRI 8 weeks −6 months after injury), where secondary injuries such as regional atrophy and microstructural alterations become apparent; and chronic (MRI more than 6 months after injury), when some recovery and/or atrophy continues, but the brain is considered to be more stable. As discussed in a prior paper,^31^ the exact boundaries were based on published data and natural break points within our datasets. Within the msTBI group, 67 scans were conducted during the acute phase, 122 during the post-acute, and 224 during the chronic phase of injury. Seven of the studies were longitudinal and five were cross-sectional, generating a total of 783 scans from 598 participants, 185 of whom had data for two timepoints.

**Table 1.** Cohort Demographics. For each cohort, the total sample size included (N) is shown, along with the number of: scans, participants with longitudinal data, msTBI and non-TBI participants, and sex. The age range (in years) and average age of cohorts are also shown with standard deviation. * indicates sites with orthopedic injury (OI) non-TBI participants versus healthy controls. UTHouston had both OI and healthy controls. ^⍭^ indicates sites with longitudinal data.

| Cohort | Total N | Total Scans | Longitudinal | msTBI | Comparison | M | F | TSI range (weeks) | Age Range | Average Age (SD) |
| --- | --- | --- | --- | --- | --- | --- | --- | --- | --- | --- |
| RAPBI<sup>†</sup> | 109 | 158 | 49 | 53 | 56 | 72 | 37 | 3.9-36.3;<br>49.2-82.7 | 8.40-19.70 | 15.61 (2.79) |
| Pilot-RAPBI<sup>†</sup> | 22 | 24 | 2 | 13 | 9 | 14 | 8 | 11.7-36.2;<br>59.1-68.6 | 12.10-18.57 | 16.14 (1.86) |
| NCH* | 53 | 53 | 0 | 29 | 24 | 37 | 16 | 58.7-424 | 8.16-16.52 | 11.80 (2.38) |
| Kennedy-Krieger<sup>†</sup> | 42 | 55 | 13 | 29 | 13 | 27 | 15 | 4.1-14.6;<br>51.9-760 | 8.12-18.98 | 14.57 (2.43) |
| LLU<sup>†</sup> | 52 | 100 | 48 | 21 | 31 | 34 | 28 | 1.0-2.6;<br>48.1-62.1 | 5.45-17.78 | 12.89 (3.45) |
| Deakin1 | 44 | 44 | 0 | 18 | 26 | 20 | 24 | 15.6-562 | 8.52-19.00 | 14.34 (2.80) |
| Deakin2 | 49 | 49 | 0 | 22 | 27 | 23 | 26 | NA | 8.52-18.97 | 14.60 (2.73) |
| BCM1*<sup>†</sup> | 99 | 134 | 35 | 50 | 49 | 70 | 28 | 11.7-28.8;<br>49.6-117.8 | 7.44-18.70 | 13.53 (2.83) |
| BCM2* | 32 | 32 | 0 | 15 | 17 | 21 | 11 | 0.14-15.4 | 10.67-19.40 | 14.56 (2.62) |
| BCM3* | 31 | 31 | 0 | 22 | 9 | 21 | 10 | 2.4-60.0 | 10.61-18.51 | 15.48 (2.34) |
| Murdoch<sup>†</sup> | 22 | 43 | 21 | 22 | 0 | 17 | 5 | 4.5-37.6;<br>99.8-124 | 5.83-16.83 | 11.13 (2.86) |
| UTHouston*<sup>†</sup> | 43 | 60 | 17 | 20 | 23 | 29 | 14 | 4.1-18.3;<br>55.8-63.0 | 8.16-16.91 | 12.70 (2.41) |
| Total | 598 | 783 | 185 | 314 | 284 | 385 | 222 |  | 5.45-19.70 | 14.05 (3.06) |

### Standard Protocol Approvals, Registrations, and Patient Consents

Original studies were approved by the individual IRBs for each respective institution. All participants provided written or verbal informed assent, while parents provided written informed consent.

### Image Acquisition, Processing, and Quality Control

Raw 3D T1-weighted MR images were processed using the ENIGMA Cerebellum Pipeline, which is based on the Automatic Cerebellum Anatomical Parcellation using U-Net with Locally Constrained Optimization (ACAPULCO; v 0.2.1) workflow.^5,35,36^ Image processing, segmentation, and quality review were conducted at the University of Utah. The cerebellum was segmented into 28 subregions (**Figure 1**). Segmentations were visually quality checked, and statistical outliers (> 3 SDs), identified for each ROI separately, **Supplementary Table 3**) were excluded. Across regions, between 7-19 outliers were removed from the 783 scans. Having an outlier volume for one region did not exclude a participant entirely, but rather just from those regional analyses. Participants whose total cerebellum volume was an outlier were removed from all analyses (N = 13). There were no significant differences in the QC fail rate between msTBI and non-TBI for any of the regions. We also checked each scan for cerebellar lesions (visible on T1-weighted scans only). These were relatively rare, with only 14% of msTBI participants having visible lesions in the cerebellum. We examined volume of the total cerebellum, corpus medullare, 5 vermal regions, and 11 lateralized lobules (averaged between left and right), for a total of 18 ROIs. We also conducted analyses for the subset of participants with longitudinal data (two timepoints) through an updated version of ACAPULCO that has been further optimized for longitudinal analysis (v 0.3.0).^5,37^

**Figure 1.**
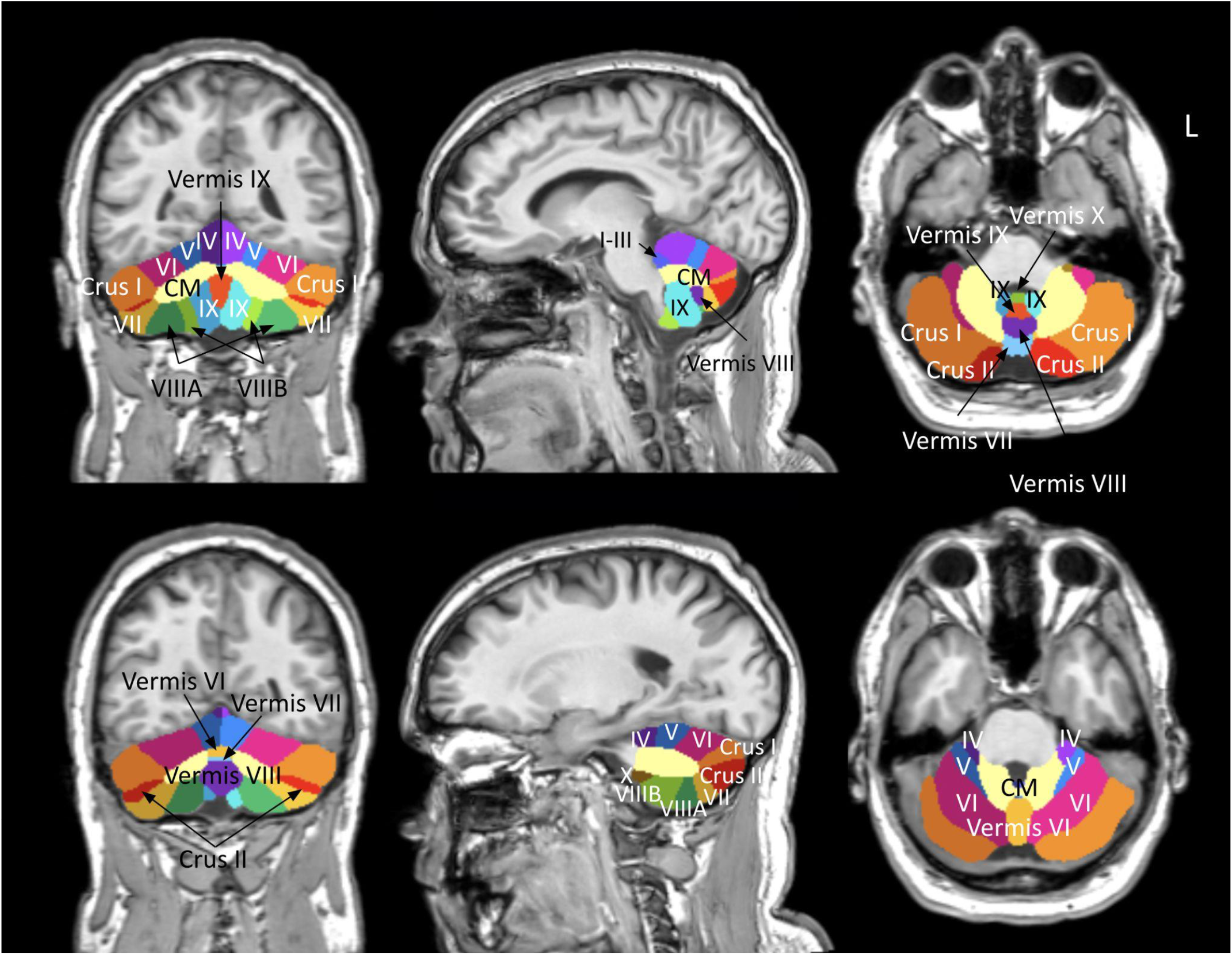
ACAPULCO segmentation. The parcellation of the cerebellum according to ACAPULCO. Images are viewed in radiology space where image left corresponds to the subject’s right side. CM=corpus medullare.

DTI data were processed as per a prior analysis and full details may be found in our previous publication.^31^ Briefly, data were processed through the ENIGMA-DTI pipeline (http://enigma.ini.usc.edu/protocols/dti-protocols/), which is a modified tract-based spatial statistics (TBSS) approach^38^ resulting in FA and other metrics averaged within a set of regions of interest (ROIs) from the Johns Hopkins University atlas. Of the 12 cohorts included here, 10 collected DTI data (parameters may be seen in **Supplementary Table 4**).

### Neurobehavioral Measures

As a secondary analysis of data that was collected through different studies, there was considerable variability in the neurobehavioral scales collected. We limited our analyses to those inventories that were collected by the most cohorts. Work is on-going in the ENIGMA Brain Injury working group to harmonize scales within the same domain.^39^

The Behavior Rating Inventory of Executive Function (BRIEF) is one of the most widely used parent/informant questionnaire measures of executive functioning in children.^40^ Parents respond to an 86-item inventory about their child’s behaviors and their responses are used to derive three age-adjusted index scores. The Behavioral Regulation Index (BRI) score measures cognitive abilities, such as inhibition, task shifting, emotional control, and self-monitoring, while the Metacognition Index (MCI) score measures initiation, working memory, planning/organizing, and task monitoring. The Global Executive Composite (GEC) is an overarching summary score of executive functioning. For each of the three subscores, a higher score indicates greater executive dysfunction. The total number of participants who completed the BRIEF was 421 (msTBI; n = 232, non-TBI; n = 189). The average, SD, and range of *T*-scores for the BRIEF measures within the msTBI group were as follows: for BRI, M = 51.2±11.4; for MCI, M = 52.8±11.3; for GEC, M = 52.4±11.4. Statistical outliers (>3 SDs) were removed (BRI = 7, MCI = 2, GEC = 4).

The Delis-Kaplan Executive Function System (D-KEFS) is a standardized battery of neuropsychological tests used to evaluate higher-order cognitive function in children and adults. The sub-test utilized in the current study was the D-KEFS Trail Making Test (TMT) conditions 3 and 4, which involve a visuo-motor sequencing task that measures processing speed (condition 3) and cognitive switching (condition 4; a measure of performance-based executive functioning). The total number of participants that completed the D-KEFS was 231 (msTBI; n = 110, non-TBI; n = 121). The average, SD and range of standardized scores for the D-KEFS measures were as follows: TMT 3, M = 9.78±9.90, range = 1-15; TMT 4, M = 9.17±5.66, range = 1-15. Higher scores indicate better performance.

### Statistical Analysis

The total number of participants was 598, with 185 participants having longitudinal data. For sites with multiple datasets, each dataset was analyzed as a separate cohort, yielding 12 cohorts in total. Linear mixed-effects models were conducted in R 3.1.3 with the *nlme* package (r-project.org/). Random effects (intercept) were used to control for site and participant ID. All analyses covaried for age, sex, and intracranial volume (ICV). We computed Cohen’s *d* effect sizes with 95% confidence intervals and unstandardized beta values for continuous predictors. Cohen’s *d* values were calculated using the following equation:

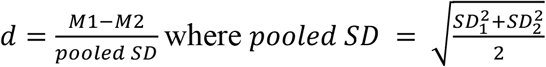

We used a modified Bonferroni correction for multiple comparisons created by Li and Ji,^41^ which yielded the effective number of independent variables (V_eff_) in our analysis as 11 and a significance threshold of p < 0.05/11 = 0.0045. A traditional Bonferroni correction was too conservative for our analysis because there are correlations between test statistics (from adjacent subregional cerebellum volumes).

We calculated corrected *p*-values using the following equation:

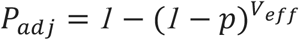

Where *p* is the unadjusted *p*-value. A flowchart showing the analyses reported below may be found in **Supplementary Figure 1**.

#### Non-linear Age Term

We first tested whether a nonlinear age term (age^2^) should be included in statistical models along with age, sex, and ICV. Age^2^ was not significant (*p*’s > 0.10), so it was not included in subsequent analyses.

#### Primary Group Comparisons

Our primary analyses compared children with a history of msTBI to non-TBI samples. In our primary group analysis, we examined group differences covarying for age, sex, and ICV including all post-injury windows. One site lacked a non-TBI group and was omitted from group analyses (Murdoch). Further, we examined differences in changes in total cerebellum volume in the subset of participants who were scanned at two timepoints and whose scans passed quality control (n = 80), covarying for scan interval and time since injury (TSI) at first scan, with site as a random effect. Within those 80 participants, the average interval between scans was 1.1 years (SD = 0.3, range = 0.7-1.9 years).

#### Secondary Group Comparisons

We conducted secondary group comparisons to characterize group effects in greater depth, including covarying for TSI and excluding patients in the acute phase when acute pathology may be influencing neuroimaging metrics. We also conducted analyses separating cohorts based on non-TBI population (HC vs. OI), separating by severity, and separating by phase of injury (acute, postacute, chronic). To ensure that cerebellar lesions were not driving our results, we conducted supplementary sensitivity/control analyses excluding any participants with cerebellar lesions visible on T1-weighted images (n = 525).

#### Exploratory Multimodal Analyses

Based on results of the primary and secondary group comparisons, we conducted an exploratory multimodal analysis in the msTBI group examining whether DTI FA predicted changes in total cerebellum volume. As detailed in our prior paper, FA was estimated and mapped onto the ENIGMA DTI template, projected onto the WM skeleton, and averaged within each of 27 partially overlapping regions of interest from the Johns Hopkins Atlas (enigma.ini.usc.edu/protocols/dti-protocols/).^31^ Lateralized ROIs were averaged. We then examined associations between FA and regional cerebellar volumes in the msTBI group collected in the same scan session (n = 252) covarying for age, sex, ICV, and GCS. Further, we explored the predictive value of FA. To this end, we included 43 participants from the msTBI group for which we had high-quality DTI data at time point 1 (acute or post-acute) and high-quality cerebellar segmentations at time points 1 and 2. With some participants missing covariates such as GCS (measure of injury severity), the final sample size for this analysis was 32. With the relatively small sample size, we consider these results exploratory.

#### Interactions

For cerebellum volumes, we examined potential interactions with group, including age and sex. Within the msTBI group, we examined potential interactions between age at injury and TSI, age at injury and GCS, and GCS and TSI.

#### Injury Variables

Using regression analysis, we examined linear relationships within the msTBI group. Covarying for age, sex, and ICV, we investigated correlations between cerebellar volume and age at injury, TSI, and GCS.

#### Data Availability

Data are available to researchers who join the working group and submit a secondary analysis proposal to the group for approval, which is granted on a cohort-by-cohort basis.

## Results

All *p*-values reported below are adjusted for multiple comparisons unless otherwise specified using the Li and Ji-modified Bonferroni approach.^41^

### Primary Group Comparisons

#### Cross-sectional

All participants (with the exception of the Murdoch cohort) and covariates age, sex, and ICV were included in this analysis, along with random effects for site and subject. Including data from all time points, participants in the msTBI group had significantly smaller volumes of the total cerebellum (*d = −*0.37, *p <* .001), corpus medullare (*d = −*0.43, *p <* .001), Crus II (*d* = −0.32, *p* < .001), Lobule VIIB (*d =* −0.25, *p* < .05), Lobule VIIIB (*d =* −0.39, *p <* .001), Vermis VII (*d = −*0.23, *p <* .005), and Vermis IX (*d =* −0.22, *p <* .05), compared to the non-TBI group. Results are summarized in **Figure 2** and **Table 2**. Removing any participants with visible cerebellar lesions yielded similar results, although the vermal group differences were no longer significant.

**Table 2:** Group Comparison.

| Region | Subregion | *p* | Adjusted *p* | Cohen's d-value | 95% CI |
| --- | --- | --- | --- | --- | --- |
| Total Volume |  | **2.1E-08** | **2.3E-07** | **-0.37** | **[-0.52,-0.22]** |
| Corpus Medullare |  | **1.7E-06** | **1.9E-05** | **-0.43** | **[-0.58,-0.28]** |
| Anterior Lobe | Lobule I-III | 0.32 | 0.98 | -0.08 | [-0.23,0.07] |
|  | Lobule IV | 0.75 | 1.0 | 0.02 | [-0.13,0.17] |
|  | Lobule V | 0.017* | 0.17 | -0.18* | [-0.33,-0.03] |
| Posterior Lobe | Lobule VI | 0.027* | 0.26 | -0.18 | [-0.34,-0.02] |
|  | Crus I | 0.011* | 0.12 | -0.23 | [-0.41,-0.05] |
|  | Crus II | **1.1E-04** | **1.2E-03** | **-0.32** | **[-0.49,-0.16]** |
|  | Lobule VIIB | **1.6E-03** | **0.018** | **-0.25** | **[-0.41,-0.10]** |
|  | Lobule VIIIA | 0.50 | 1.0 | -0.06 | [-0.22,0.11] |
|  | Lobule VIIIB | **5.8E-06** | **6.4E-05** | **-0.39** | **[-0.56,-0.22]** |
|  | Lobule IX | 0.033* | 0.31 | -0.17* | [-0.33,-0.01] |
| Flocculonodular Lobe | Lobule X | 0.98 | 1.0 | 0.00 | [-0.15,0.15] |
| Vermis | Vermis X | 0.013* | 0.13 | -0.19* | [-0.34,-0.04] |
|  | Vermis VI | 0.41 | 1.0 | 0.06 | [-0.09,0.21] |
|  | Vermis VII | **2.8E-03** | **3.0E-02** | **-0.23** | **[-0.38,-0.08]** |
|  | Vermis VIII | 0.12 | 0.76 | -0.12 | [-0.27,0.03] |
|  | Vermis IX | **0.0044** | **0.047** | **-0.22** | **[-0.37,-0.07]** |

The p-values, adjusted p-values, Cohen d-values, and the 95% confidence intervals for d-values are shown for the group comparison.
Bolded values are significant (based on Li and Ji adjusted Bonferroni correction)
\*Results are significant at $p < 0.05$

**Figure 2.**
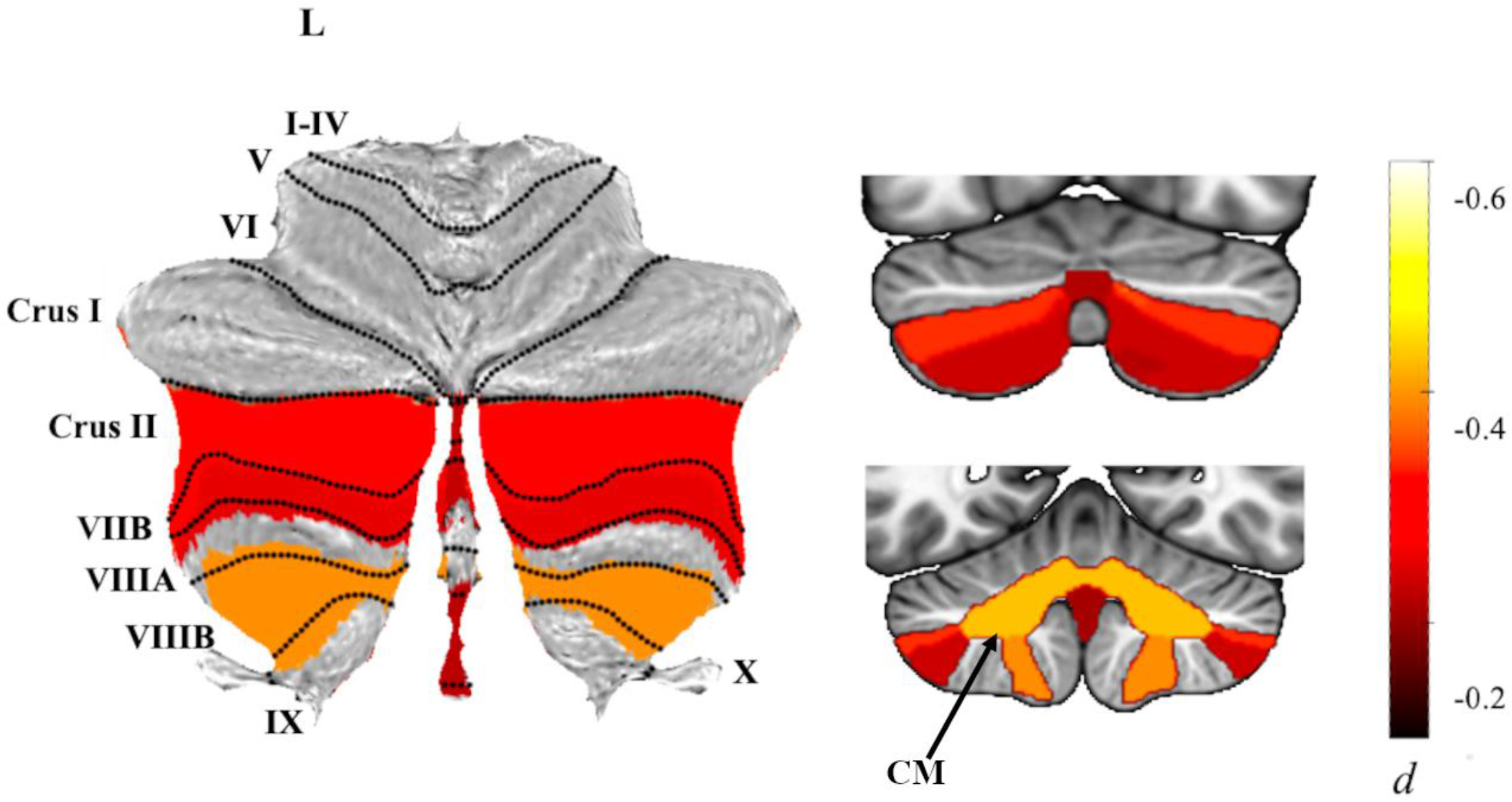
Group comparison. Atlas-based effect size (Cohen’s *d*) maps and MNI-based coronal slices (top: y=-72; bottom y=-54) of the significant between-group differences for children with msTBI vs. controls.

We separated participants based on TSI, comparing the msTBI and non-TBI groups in the acute phase, post-acute phase, and chronic phase (**Supplementary Table 5)**. We found that significant group differences were predominantly driven by participants in the chronic phase, with no significant differences surviving multiple comparisons correction in the post-acute phase, and a significantly smaller Vermis VII in the acute phase.

Additionally, we categorized participants based on severity level (complicated mild TBI, moderate TBI, severe TBI) and found group differences for severe TBI in volumes of the total cerebellum (*d=*-0.33, *p*<.001) corpus medullare (*d* = −0.42, *p <* .001), Lobule V (*d =* −0.26, *p <* .001), Crus I (*d* = −0.33, *p* < .005) and Lobule VIIIB (*d =* −0.38, *p <* .001), and significant differences for moderate TBI in volumes of the corpus medullare (*d =* −0.30, *p* < .001) and Lobule VIIB (*d =* −0.38, *p* < .001) (**Supplementary Table 6**). There were no significant differences between the complicated mild TBI and non-TBI groups in subregional cerebellum volumes.

The above severity and chronicity analyses were run separately for six different comparisons (three for each). To visualize effects, we also ran these combined, for nine different comparisons. These results are tiled by severity and chronicity in **Figure 3**.

**Figure 3.**
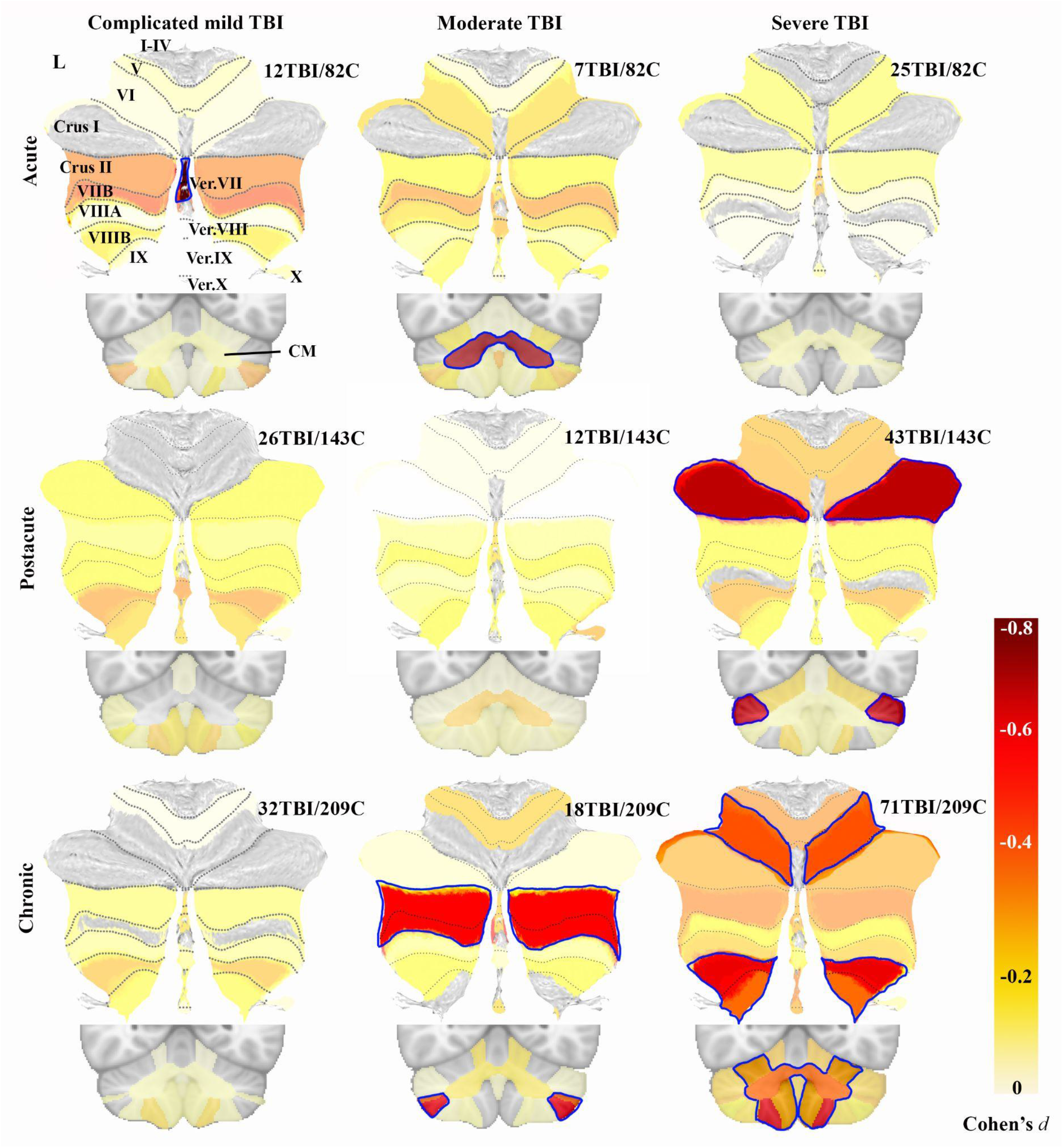
Severity and chronicity analyses. Atlas-based effect size (Cohen’s *d*) maps and MNI-based coronal slices are shown for group comparisons separated by severity (columns) and chronicity (rows). Lobules and vermal regions (Ver.VII-X) are labeled in the top left on the SUIT flatmap. The corpus medullare (CM) is shown in the coronal slices. The color corresponds to the effect size, according to the colorbar, with dark red for the largest effect sizes. Non-significant effect sizes are shown at 50% opacity, while significant ones are not opaque and outlined in blue. The number of msTBI and non-TBI participants (C) are shown for each comparison. No participant was included twice in any of the nine sub-analyses, but non-TBI participants were included across multiple comparisons. Only negative effect sizes are shown, positive effect sizes were not significant and are not included.

#### Longitudinal

When examining longitudinal changes in total cerebellum volume for the subset of participants with longitudinal data (n = 75), we found significantly slower growth and/or greater decreases in volume in the msTBI group than the non-TBI group (*d* = −0.55, *p* = .021; **Figure 4**). This effect persisted when we excluded msTBI participants with cerebellar lesions (n = 73, *d* = −0.47, *p* = .024), and when covarying for changes in total brain volume (*d* = −0.62, *p* = .01). Total cerebellum volume decreased for roughly half of the msTBI participants, generally those injured at a younger age. Within the msTBI group, changes in total cerebellum volume were positively associated with age at injury (*b* = 0.0052, *p* = .01, **Supplementary Figure 2**) and not associated with participant sex or GCS.

**Figure 4.**
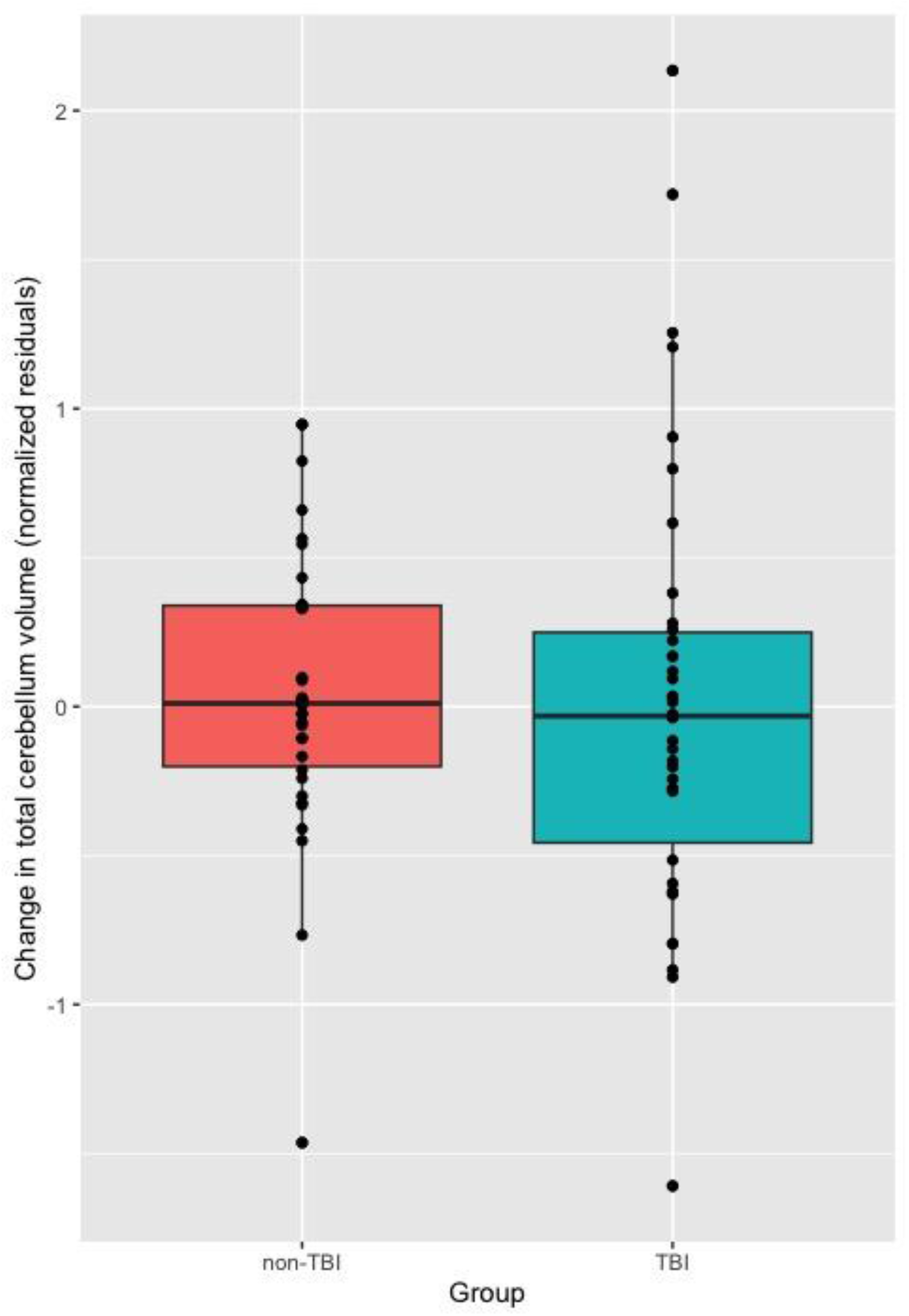
Group differences in longitudinal changes in cerebellum volume. Longitudinal changes in total cerebellum volume are shown for the subset of participants with high-quality longitudinal data, outliers (3SD) removed (N=72). The boxplot shows the percent volume change (normalized residuals) for the non-TBI group (pink, N=34), and the msTBI group (blue, N=38).

### Secondary Group Comparisons

We conducted a number of *post hoc* tests to examine potential confounds. We tested several additional models covarying for TSI, excluding participants in the acute phase (< 7 weeks post-injury), or both excluding acute and covarying for TSI. Results were consistent with our primary model (**Supplementary Table 7**). Further, we examined group differences separately for sites that had HC versus those with OI controls (**Supplementary Table 8**). We found significant differences only with the HC comparison, while differences between msTBI and OI were no longer significant. However, ADHD is a common pre-/comorbidity of TBI^42^ and is associated with cerebellar changes,^35^ so we also examined only HC and OI sites that excluded children with ADHD. When excluding HC and OI cohorts that did not exclude children with ADHD, the HC and OI results were very similar (**Supplementary Table 8**).

### Exploratory Multimodal MRI Analyses

As the multimodal analyses were exploratory, we used an uncorrected threshold of *p*<.05 and uncorrected *p*-values are reported below. In the msTBI group, there were significant correlations between total cerebellum volume and FA for a number of white matter ROIs cross-sectionally (**Supplementary Table 9**). This relation was significant when covarying for GCS, suggesting that the relation between cerebellar volume and FA in the cerebrum is (at least partly) independent of the initial impact of TBI severity. We found significant associations between FA at baseline with longitudinal changes in total cerebellum volume, with higher FA in the body of the corpus callosum, cingulum, hippocampal cingulum, corona radiata, fornix, inferior cerebellar peduncle, posterior thalamic radiation, retrolenticular limb of internal capsule, superior longitudinal fasciculus, sagittal stratum, and tapetum associated with increases in cerebellum volume in the msTBI group (**Supplementary Table 9** and **Supplementary Figure 3**). Higher average FA across the white matter was also associated with changes in cerebellum volume. For this analysis we covaried for interval, TSI (first scan), GCS, and percent change in ICV, indicating that the associations between baseline FA and secondary cerebellar changes were not due to injury severity or overall atrophy. The only significant cross-sectional or longitudinal associations between FA and total cerebellum volume in the non-TBI group were the cerebellar peduncles.

### Interactions

The rest of the analyses were corrected for multiple comparisons as reported above, with corrected *p*-values reported. We did not find any significant interactions between group and age or group and sex. There were also no significant interactions within the msTBI group between age at injury and TSI or between age at injury and GCS when covarying for age at scan (**Supplementary Table 10**). We found a significant interaction between TSI and GCS for total cerebellum volume (*b* = 6.3, *p* < .001; **Supplementary Figure 4**), with participants with higher GCS scores (milder injuries) showing volume increases with further time since injury and those with lower GCS scores showing decreases in volume over time.

### Injury Variables

Within the msTBI group, we did not find any significant associations with TSI, GCS, or age at injury (controlling for age at scan).

### Executive Functioning

Executive functioning measures were only examined for associations with cerebellar volume in the msTBI group across all available time points. Lower cerebellar volume was significantly associated with more parent-reported problems in executive functioning within the msTBI group. There was a significant negative association between total cerebellum volume and BRIEF MCI (*b* = −202, *p <* .05) and GEC (*b =* −209, *p <* .001) scores, such that smaller cerebellar volumes were related to greater executive dysfunction. No significant associations were observed for the BRIEF BRI score. Results are shown in **Table 3**. Lower cerebellar volume was also associated with slower processing speed in participants with msTBI. For the D-KEFS TMT, there was a significant association between the condition 3 score and the volume of Lobule I-III (*b =* 4.0, *p =* 0.022). Results are summarized in **Supplementary Table 11**.

**Table 3:**
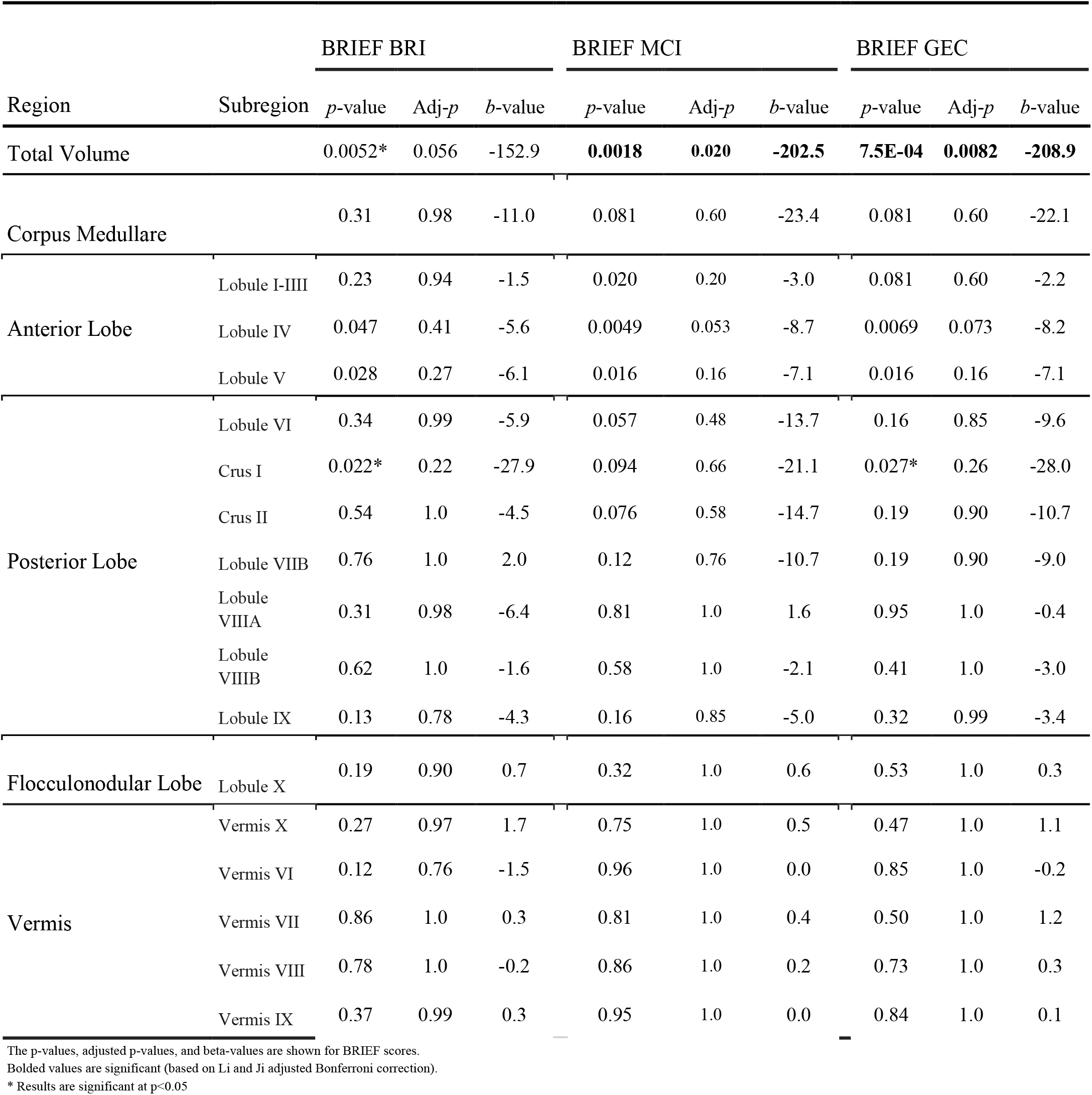
BRIEF Scores.

## Discussion

Using a multi-site, multimodal, longitudinal dataset compiled by the ENIGMA Pediatric Moderate/Severe TBI Working Group, we analyzed regional cerebellum volume in a cohort of 598 children/adolescents with or without msTBI and examined associations with executive functioning. This is the largest published study to date using structural MRI in children with msTBI. We hypothesized that participants with msTBI would have, on average, smaller cerebellar volume than the non-TBI group and that these volumetric alterations would be most prominent in patients greater than six months post-injury. Our results support these hypotheses, with participants in the msTBI group demonstrating smaller total cerebellum volume, as well as smaller volume in six subregions in the posterior lobe and vermis. These volume reductions were shown to be most prominent in patients with more severe injuries and those who were at least six months post-injury, suggesting that volumetric changes in the cerebellum may be due to a secondary injury process. This secondary injury hypothesis was substantiated with longitudinal analyses in a subset of the data incorporating multimodal MRI. Lastly, we hypothesized that volume alterations would be associated with poorer executive functioning.

### Cerebellum Development

Prior studies have shown complex developmental trajectories of the cerebellum, with peak maturation trending first from the vermis and Lobule IX at age 5, to the anterior lobe between ages 12 and 16, and to the posterior lobe between adolescence and early adulthood (other than Lobule IX). Based on these trajectories, it is likely that within our sample of participants, posterior cerebellar regions were still undergoing structural maturation at the time of injury and MRI scan (mean age at injury = 13.0±3.6 years, mean age at scan = 14.0±3.1 years). As we found the majority of significant volumetric differences between the msTBI and non-TBI groups in the posterior lobe, our results suggest that the immaturity of subregions in the cerebellum at the time of injury may contribute to their vulnerability. Plasticity during development is a double-edged sword, potentially leading to faster recovery, but also increasing susceptibility to disruption.^43^ Development may not be the major driving factor here, however, as it is possible that the posterior cerebellum is particularly susceptible to disruption, perhaps due to its structural and functional connectivity with the frontal cortex.^44–46^ Two recent examinations in ENIGMA working groups have shown deficits related to PTSD and epilepsy, primarily in the posterior lobe.^36,47^ In contrast to these previous investigations involving adult clinical populations, our results offer novel findings from a relatively large, pediatric brain injury population. Further analyses with expanded age ranges and multi-modal MRI data may be able to disentangle these potential sources of susceptibility.

### Potential Sources of Cerebellar Vulnerability

The same fronto-cerebellar networks that explain the cerebellum’s role in cognitive function may also be a source of indirect injury to this region. In the absence of direct injury, atrophy in the cerebellum may be due to a secondary injury process such as connectomal diaschisis. We found alterations in the gray and white matter of the posterior lobule, with the largest effect size in the *corpus medullare* (*d* = 0.43). This is where the deep cerebellar nuclei, the terminus for many cortical projections, are located. With our data, we cannot resolve the specific nuclei and the fibers terminating there, but this could indicate that the connectivity of the cerebellum underlies the structural vulnerability. If this was the case, we would expect that deficits in volume would not be seen until the chronic phase of injury. Separating our sample by acute, post-acute, and chronic timepoints, we found significant differences in volume between the msTBI and non-TBI groups in the chronic phase of injury for total cerebellum volume and six cerebellar subregions. The only significantly lower subregional volume for participants in the acute phase of injury was in Vermis VII, and there were none in the post-acute phase of injury after correcting for multiple comparisons. This suggests that atrophy in the cerebellum may occur months after the initial insult. We substantiated these findings by conducting a secondary analysis on participants with longitudinal data and found greater total cerebellum volume decreases in the msTBI group. Additionally, in a longitudinal analysis of the subset of participants with DTI at time point 1 and cerebellar segmentations at time points 1 and 2 (n = 32), we found further evidence supporting that volumetric reductions in the cerebellum may be related to prior white matter alterations. Lower FA of multiple central white matter regions was associated with slower growth and/or greater atrophy in the cerebellum. These results were significant when controlling for injury severity and change in total brain volume, indicating that they are not simply linked through general neuropathology post-injury. These associations between FA and cerebellum volume were also only present in the non-TBI group in the peduncles, supporting the interpretation that these associations are largely unique to the context of injury. This supports the hypothesis that volumetric deficits in the cerebellum are due to a secondary disease process such as connectomal diaschisis, not acute injury. The link between the specific white matter regions we report and other sources of acute pathology, such as lesions, is not clear, and the full extent of the interconnected structural and functional network that leads to cerebellar vulnerability has yet to be shown.

### Cerebellum and Cognition

Our results suggest that within the msTBI group, the volume of the total cerebellum was associated with lower parent-reported executive functioning scores, as measured by the BRIEF MCI and GEC scores. Slower processing speed (D-KEFS TMT 3) was associated with the volume of Lobule I-III. Cerebellar support of cognitive function may be due in part to fronto-cerebellar circuits. Studies have demonstrated multiple segregated fronto-cerebellar networks connecting the cerebellum to frontal regions such as the dorsolateral, medial, and anterior prefrontal cortex,^48^ all of which are distinctly involved in cognitive function. The posterior lobe of the cerebellum, where we observed the greatest volumetric deficits between the msTBI and non-TBI groups, is of particular interest because it is notably larger in humans and apes compared to other mammals^49^ and has been shown during phylogenetic expansion to mirror the frontal cortex.^45,46^ Recent functional studies have observed that the majority of the posterior cerebellum is associated with cognitive regions of the neocortex, while anterior and vermal regions demonstrate less involvement in cognitive function.^44^ Lesion analyses have revealed functional divisions in the cerebellum-anterior lobe lesions leading to motor syndromes, posterior lobe lesions leading to cognitive impairments, and vermis lesions leading to neuropsychiatric symptoms.^50^ In truth, the divisions are not quite so absolute, with the inferior posterior cerebellum (lobules VIIIA-IX) also involved in motor planning.^20^

### Potential Confounds

One important confounding variable is the occurrence of pre-existing psychiatric disorders within our participant sample. In particular, ADHD is a significant risk factor for TBI^51^ and also shows associations with smaller cerebellum volume.^52^ We thus conducted a secondary analysis only using data from sites that excluded participants with ADHD and found largely consistent results. Additionally, it has been shown that cerebellar differences in participants with ADHD are due to delayed developmental trajectories that demonstrate a pattern of normalization with age - cerebellar volumes demonstrate greater disparities at a younger age and eventually match healthy controls in late adolescence.^53^ As such, ADHD-related volume deficits would be expected throughout all phases of injury in our sample.^35^ However, our results demonstrated significantly greater deficits in subregional cerebellar volume in the chronic phase of injury, suggesting that ADHD was not a confounding factor.

### Limitations

Important limitations of our study are the differences among recruitment criteria, scan parameters, and collected measures between sites. Despite our large sample size, these inconsistencies limit the power of some of our analyses. For example, the broad variability across sites at the time of testing/scanning post-injury may limit our results, as the first year after injury is particularly dynamic.^31^ We established post-injury intervals and conducted analyses within phase to better understand this phenomenon. However, this approach is inherently flawed as the physiological changes that occur are along a continuum, rather than in discrete periods. Further, there were differences in the numbers of participants within the defined phases. Our scan totals for acute, postacute, and chronic patients were 115, 228, and 394, respectively. Similarly, our longitudinal analysis consisted of only 80 participants. However, while the multi-site design of this study led to variability, it also allowed us to obtain the largest sample of pediatric msTBI to date, demonstrating the utility of ENIGMA to generate and answer hypotheses that would otherwise be inaccessible. Another limitation of our study is the inability to control for premorbid psychiatric, behavioral, or neurological factors.^54^ Many of the resulting symptoms measured in our analyses are also risk factors for TBI and could have existed pre-injury. The most significant pre-injury condition in regard to the cerebellum is ADHD, but this was controlled for as described above. Finally, while we were able to incorporate multimodal data, TBSS is an ROI-based approach, not tractography, limiting our ability to fully attribute our results to specific tracts. To more surely elucidate this as a mechanism of injury to the cerebellum, further mapping of the structural connectome using fiber tracking may be useful.

### Conclusion

In a sample of nearly 600 participants with or without pediatric msTBI from the ENIGMA Pediatric msTBI working group, cerebellum volume was significantly smaller in patients with TBI. Subregional volumetric deficits were most pronounced in the later-developing posterior cerebellum, suggesting an increased vulnerability to injury in developing regions. Consistent with recent findings establishing the cerebellum’s role in cognition, volumetric alterations were associated with poorer executive functioning. Our longitudinal and multimodal results suggest that indirect cerebellar injury may be partially influenced by injury-related disruptions in cerebral white matter microstructure. Future research incorporating injury regions and fronto-cerebellar circuits is required to further understand the mechanisms underlying cerebellar injury after TBI in children.

## Supporting information

STROBE

## Acknowledgments

We gratefully acknowledge the contributions of our undergraduate research assistants in reviewing the cerebellar parcellations: Teric Abunuwara, Lauren Christensen, Tanner Guderian, Karissa Liu, Lindsey Rose Melby, Osvaldo Miranda, and Michael Pink.

