## Supplement for "Multimodal Analysis of Secondary Cerebellar Alterations after Pediatric Traumatic Brain Injury"

**Supplementary Materials**

**Supplementary Table 1. Cohort details**. For each cohort, the recruitment location is shown, along with inclusion and exclusion criteria.

**Supplementary Table 2. Scan and protocol details.** For each cohort the scanner manufacturer and model are listed, along with field strength and voxel size (in mm).

**Supplementary Table 3. Quality control exclusions by ROI and site.** Outlier / visual QC exclusion counts by cerebellar subregion per site.

**Supplementary Table 4. DTI protocol details.**

**Supplementary Figure 1. Flowchart of analyses.**

**Supplementary Table 5. Phase of injury:** Effects of phase of injury (Acute: <7 weeks, Post Acute: 7 weeks-6 months, Chronic: >6 months) on cerebellum volumes.

**Supplementary Table 6. Severity comparisons.** Effects of injury severity as rated by the GCS on cerebellum volumes.

**Supplementary Figure 2.** Association between age at injury and percent change in total cerebellum volume for the subset of TBI participants with longitudinal data available.

**Supplementary Table 7. Supplemental group comparisons.** Results are shown for the main group comparison 1) excluding participants within 7 weeks of injury, 2) excluding participants within 7 weeks of injury and covarying for time since injury (TSI), and 3) covarying for TSI.

**Supplementary Table 8. Site type:** Effects of site type ( OI comparisons or healthy controls), and presence of ADHD in cohort on cerebellum volumes.

**Supplementary Table 9.** Cross-sectional and longitudinal associations between FA and total cerebellum volume. Longitudinal associations are between FA at first scan and changes in total cerebellum volume. FA=fractional anisotropy, ACR=anterior corona radiata, ALIC=anterior limb of internal capsule, BCC=body of corpus callosum, CC=corpus callosum, CGC=cingulum, CGH=hippocampal cingulum, CR=corona radiata, CST=corticospinal tract, EC=external capsule, FX=fornix, FXST=fornix stria terminalis, GCC=genu of corpus callosum, IC=internal capsule, PCR=posterior corona radiata, PLIC=posterior limb of internal capsule, PTR=posterior thalamic radiation, RLIC=retrolenticular limb of internal capsule, SCC=splenium of corpus callosum, SCR=superior corona radiata, SFO=superior fronto-occipital fasciculus, SLF=superior longitudinal fasciculus, SS=sagittal stratum, TAP=tapetum, UNC=uncinate. Significant associations are **bolded**.

**Supplementary Figure 3.** White matter regions associated with longitudinal changes in total cerebellum volume. Red=body of corpus callosum, green=cingulum, dark blue=hippocampal cingulum, light blue=retrolenticular limb of internal capsule, yellow=sagittal stratum, orange=fornix stria terminalis, white=tapetum.

**Supplementary Table 10. Interactions.** Interaction effects between relevant independent variables (group and age, group and sex, age at injury and time since injury, age at injury and GCS score, and time since injury and GCS score).

**Supplementary Figure 4. Interaction between time since injury and injury severity.** The interaction between time since injury in weeks and Glasgow Coma Scale is shown for total cerebellum volume (normalized residuals accounting for age, sex, ICV, and random effects of site and subject). TSI is truncated at 4 years. Light blue dots and dotted line are for participants with GCS in the bottom tertile (most severe), dark blue dots and solid line are for participants with GCS in the top tertile (least severe), with the middle blue dots and dashed line for participants in the middle tertile.

**Supplementary Table 11**: D-KEFS Trail Making Test Scores

**Supplementary Table 1. Cohort details**. For each cohort, the recruitment location is shown, along with inclusion and exclusion criteria. * = cohorts with OI controls.

| **Cohort** | **Recruitment** | **Inclusion criteria** | **Exclusion criteria** |
| --- | --- | --- | --- |
| RAPBI | Los Angeles County Pediatric Intensive Care Units (PICUs). Physicians identified potential participants who were contacted by a study representative. Controls recruited from community. | 1) For TBI group: non-penetrating msTBI (moderate-severe) (intake or post-resuscitation Glasgow Coma Scale (GCS) score between 3 and 12); 2) 8-19 years of age; 3) right-handed; 4) normal visual acuity or vision corrected with contact lenses/eyeglasses; and 5) English skills sufficient to understand instructions and be familiar with common words (the neuropsychological tests used in this study presume competence in English). | 1) history of neurological illness, such as prior msTBI, brain tumor or severe seizures; 2) motor deficits that prevent the subject from being examined in an MR scanner (e.g., spasms, movement disorder); 3) history of psychosis, ADHD, Tourette’s Disorder, learning disability, mental retardation, autism or substance abuse. These conditions are associated with cognitive impairments that might overlap with those caused by TBI. MRI contraindication was also an exclusion criterion. |
| Pilot-RAPBI | Los Angeles County Pediatric Intensive Care Units (PICUs). Physicians identified potential participants who were contacted by a study representative. Controls recruited from the community. | 1) For TBI group: non-penetrating msTBI (moderate-severe) (intake or post-resuscitation Glasgow Coma Scale (GCS) score between 3 and 12); 2) 8-19 years of age; 3) right-handed; 4) normal visual acuity or vision corrected with contact lenses/eyeglasses; and 5) English skills sufficient to understand instructions and be familiar with common words (the neuropsychological tests used in this study presume competence in English). | 1) history of neurological illness, such as prior msTBI, brain tumor or severe seizures; 2) motor deficits that prevent the subject from being examined in an MR scanner (e.g., spasms, movement disorder); 3) history of psychosis, ADHD, Tourette’s Disorder, learning disability, mental retardation, autism or substance abuse. These conditions are associated with cognitive impairments that might overlap with those caused by TBI. MRI contraindication was also an exclusion criterion. |
| NCH* | TBI and trauma controls were recruited by mail from institutional trauma registries in Ohio. Interested participants were screened by phone. | 1) hospitalized for at least one night for moderate to severe TBI (post-resuscitation GCS score between 3 and 12) or fracture (control OI group); 2) 8-15 years of age; 3) injured 1-4 years prior. | 1) a history of previous TBI requiring medical treatment (i.e., prior to the target injury); 2) premorbid neurological disorder or mental retardation, or full-time special education placement at school; 3) injury as a result of child abuse or assault; 4) a history of severe psychiatric disorder requiring hospitalization; 5) sensory or motor impairment that precludes completion of study measures; 6) primary language other than English; 7) any contraindication to MRI; 8) refusal by the child’s school to participate in the school visit. |
| Kennedy-Krieger | Potential participants with TBI were identified from current or historical inpatient or outpatient rehabilitation care at Kennedy Krieger Institute. Controls recruited through word of mouth and community postings. | 1) aged 8-18 years of age at time of all study visits; 2) Child and parent are conversant in English, as determined by their ability to understand and participate in English conversations to review the screening form (parent) and consent form (parent and child) 3) For TBI group: moderate or severe traumatic brain injury as defined by first Glasgow Coma Score of 12 or lower upon admission to first emergency room OR post-traumatic amnesia lasting longer than one hour OR alteration in consciousness lasting longer than 15 minutes OR injury-related intracranial abnormality on brain CT or MRI. | 1) children in foster care, 2) penetrating TBI or open TBI (evidenced by dural tear), 3) inability to complete laboratory tasks due to cognitive or motor deficits or uncorrected visual impairment (including red/green colorblindness), 4) ongoing post-traumatic amnesia at the time of first study evaluation or 5) pre-injury diagnosis of mental retardation, psychiatric disorder, or developmental disorder other than ADHD with or without Oppositional Defiant Disorder (ODD). MRI contraindication was also an exclusion criterion. |
| LLU | TBI patients were recruited from hospitals. Control participants were recruited from Loma Linda University Health System pediatric clinics. | 1) age between 4 and 18 years at the time of injury; 2) absence of previous brain injury, neurological disorders, drug or alcohol abuse, or MRI contraindications, including dental braces; 3) moderate-to-severe TBI with Glasgow Coma Scale (GCS) scores between 3 and 12 or complicated mild (cMild) TBI (GCS 13–15) if hemorrhage was detected on an acute brain imaging (CT) exam. Control subjects who met criteria 1 and 2 were recruited from our pediatric clinics and were scanned without sedation. | 1) MRI contraindications; 2) neurological disorders; 3) previous brain trauma or alcohol or drug abuse, 4) history of significant cognitive deficit (i.e. pervasive developmental disorders (PDD)) |
| Deakin University (1) |  | All moderate to severe TBI patients were assessed at least six months post-injury, when neurological recovery was stabilized. Participants were excluded  if they had pre-existing developmental or intellectual disabilities, a progressive disease, or were taking medication. | All control subjects were screened to ensure that they had no history of neurological damage. |
| Deakin University (2) | The participants were part of a larger-scale  cognitive training study in pediatric TBI | Inclusion criteria for patients were as follows: (1) Age at injury: 10–17 years; (2) Injuries classified as moderate to severe using the Mayo Classification System; and (3) In the chronic stage of injury at the time of assessment (1–5 years post injury). All participants met magnetic resonance imaging (MRI) safety criteria (i.e., no metal implants or orthodontic braces) and patients with a recurrent TBI or significant neurological illness were excluded. | Typically developing children were recruited via social networks of researchers to obtain gender- and age-matched (maximum of 6 months) controls for each TBI patient. |
| BCM (all)* | TBI and trauma control participants were recruited from Level 1 trauma centers in Houston, Dallas, and Miami. | 1) for TBI group: non-penetrating complicated mild or msTBI (moderate-severe) (post-resuscitation GCS score between 3 and 12); 2) 10-18 years of age; 3) right-handed; 4) normal visual acuity or vision corrected with contact lenses/eyeglasses; and 5) fluent in English or Spanish (the neuropsychological tests used in this study were translated into Spanish and administered by bilingual examiners). | 1) history of neurological illness, such as prior msTBI, brain tumor or severe seizures; 2) motor deficits that prevent the subject from being examined; 3) history of psychosis, Tourette’s Disorder, learning disability, mental retardation, autism or substance abuse. These conditions are associated with cognitive impairments that might overlap with those caused by TBI. Participants with contraindications to undergoing an MRI scan were excluded. |
| Murdoch | Children with TBI were enrolled in the study at the time of injury and represented consecutive  admissions to a Level 1 trauma center and ED in Melbourne, Australia | (1) documented evidence of accidental TBI, including a period of altered consciousness or presence of at least two postconcussive symptoms; (2) medical records sufficiently detailed to determine injury severity, including the Glasgow Coma Scale (GCS); and (3) child and at least one parent  fluent in English. | (1) non-accidental head injuries; (2) parent-reported history of congenital, developmental, or psychiatric condition, including history of ADHD, ASD, and Specific Learning Disorder; (3) previous TBI based on parent-report. |
| UTHouston | TBI and trauma control participants were recruited from Level 1 trauma center and ED in Houston | Inclusion criteria for TBI, OI, and healthy groups included age at injury between 8 and 15 years, proficiency in English and parent proficient in English or Spanish, residence within 125 miles, no history of prior treated TBI, no preinjury history of major neuropsychiatric disorder, no prior hospitalizations for anxiety or depression, and no history of type 1 or type 2 diabetes. Injuries were sustained in vehicle incidents. | 1) prior history of major neuropsychiatric disorder that would complicate assessment of the impact of injury on behavioral outcomes; 2) metabolic disorder; and 3) prior medically attended TBI |

**Supplementary Table 2. Scanner details.** For each cohort the scanner manufacturer and model are listed, along with field strength and voxel size (in mm).

| **Cohort** | **Scanner** | **Field strength (T)** | **Voxel size (mm)** |
| --- | --- | --- | --- |
| RAPBI | Siemens TrioTim | 3T | 1x1x1 |
| Pilot-RAPBI | Siemens Sonata | 1.5T | 1x1x1 |
| NCH | Siemens Prisma | 3T | 1x1x1 |
| Kennedy Krieger | Philips | 3T | 1x1x1 |
| LLU | Siemens TrioTim | 3T | 1x1x1 |
| Deakin1 | Siemens Magnetom Trio | 3T | 1x1x1 |
| Deakin2 | Philips Intera | 3T | 1x1x1.1 |
| BCM1 | Philips Intera or Achieva | 1.5T | 1x1x1 |
| BCM2 | Philips Intera | 3T | 1x1x1 |
| BCM3 | Philips Achieva | 3T | 1x1x1 |
| Murdoch | Siemens TrioTim | 3T | 1x0.5x0.5 |
| UTHouston | Philips | 3T | 1x1x1 |

| **Supplementary Table 3**: **Quality control exclusions by ROI and site.** Outlier / visual QC exclusion counts by cerebellar subregion per site. | | | | | | | | | | | | | | |
| --- | --- | --- | --- | --- | --- | --- | --- | --- | --- | --- | --- | --- | --- | --- |
| Region | Subregion | RAPBI | Pilot-  RAPBI | NCH | Kennedy-Krieger | LLU | Deakin1 | Deakin2 | BCM1 | BCM2 | BCM3 | Murdoch | UTHouston | Total |
| Total Volume |  | 5 | 3 | 0 | 5 | 4 | 1 | 4 | 18 | 1 | 7 | 1 | 0 | 49 |
| Corpus Medullare |  | 6 | 3 | 1 | 7 | 7 | 3 | 1 | 22 | 0 | 8 | 1 | 0 | 59 |
|  | Lobule I.III | 6 | 3 | 1 | 5 | 4 | 1 | 3 | 20 | 1 | 8 | 1 | 1 | 54 |
| Anterior Lobe | Lobule IV | 9 | 4 | 1 | 4 | 8 | 1 | 2 | 48 | 1 | 9 | 2 | 1 | 90 |
|  | Lobule V | 9 | 4 | 1 | 5 | 5 | 2 | 2 | 57 | 2 | 8 | 0 | 1 | 96 |
|  | Crus I | 47 | 22 | 6 | 3 | 14 | 9 | 22 | 105 | 3 | 10 | 24 | 3 | 268 |
|  | Crus II | 20 | 16 | 8 | 7 | 10 | 5 | 9 | 52 | 4 | 8 | 2 | 1 | 142 |
| Posterior Lobe | Lobule VI | 14 | 6 | 9 | 5 | 4 | 2 | 3 | 67 | 1 | 8 | 2 | 0 | 121 |
|  | Lobule VII | 18 | 8 | 16 | 4 | 7 | 2 | 4 | 45 | 1 | 9 | 2 | 1 | 117 |
|  | Lobule VIIIA | 16 | 8 | 11 | 5 | 10 | 3 | 5 | 75 | 1 | 8 | 3 | 8 | 153 |
|  | Lobule VIIIB | 11 | 9 | 6 | 5 | 12 | 2 | 4 | 63 | 2 | 8 | 3 | 1 | 126 |
|  | Lobule IX | 9 | 5 | 2 | 4 | 7 | 1 | 1 | 52 | 2 | 9 | 2 | 1 | 95 |
| Flocculonodular Lobe | Lobule X | 6 | 4 | 1 | 4 | 5 | 1 | 1 | 20 | 1 | 8 | 1 | 0 | 52 |
|  | Vermis X | 6 | 3 | 0 | 5 | 6 | 1 | 1 | 21 | 1 | 8 | 1 | 0 | 53 |
|  | Vermis VI | 5 | 3 | 1 | 7 | 6 | 1 | 1 | 19 | 1 | 8 | 1 | 0 | 53 |
| Vermis | Vermis VII | 8 | 3 | 0 | 4 | 5 | 2 | 1 | 21 | 2 | 9 | 2 | 1 | 58 |
|  | Vermis VIII | 5 | 3 | 1 | 5 | 5 | 1 | 1 | 19 | 1 | 8 | 1 | 0 | 50 |
|  | Vermis IX | 6 | 3 | 0 | 6 | 5 | 1 | 1 | 23 | 1 | 8 | 3 | 1 | 58 |

**Supplementary Table 4. DTI protocol details.**

| **Cohort** | **Voxel size (mm)** | **Number of gradient directions and b-value (s/mm^2^)** | **No. b0 volumes** |
| --- | --- | --- | --- |
| RAPBI | 2x2x2 | 64 at b=1000 | 8 |
| Pilot RAPBI | 2.5x2.5x2.5 | 30 at b=1000 | 5 |
| NCH | 2x2x2 | 30 at b=700 | 1 |
| Kennedy Krieger | 0.83x0.83x2.2 | 32 at b=700 | 1 |
| Loma Linda University | 1.2x1.2x3.9 | 60 at b=1000 | 2 |
| Deakin-1 | 2.2x2.2x2.2 | 64 at b=1000 | 1 |
| BCM1 | 2.7x2.7x2.7 | 15 at b=860 | 1 |
| BCM2 | 1.75x1.75x2 | 32 at b=1000 | 1 |
| BCM3 | 1.75x1.75x2 | 32 at b=1000 | 1 |

**Supplementary Figure 1. Flowchart of analyses.**

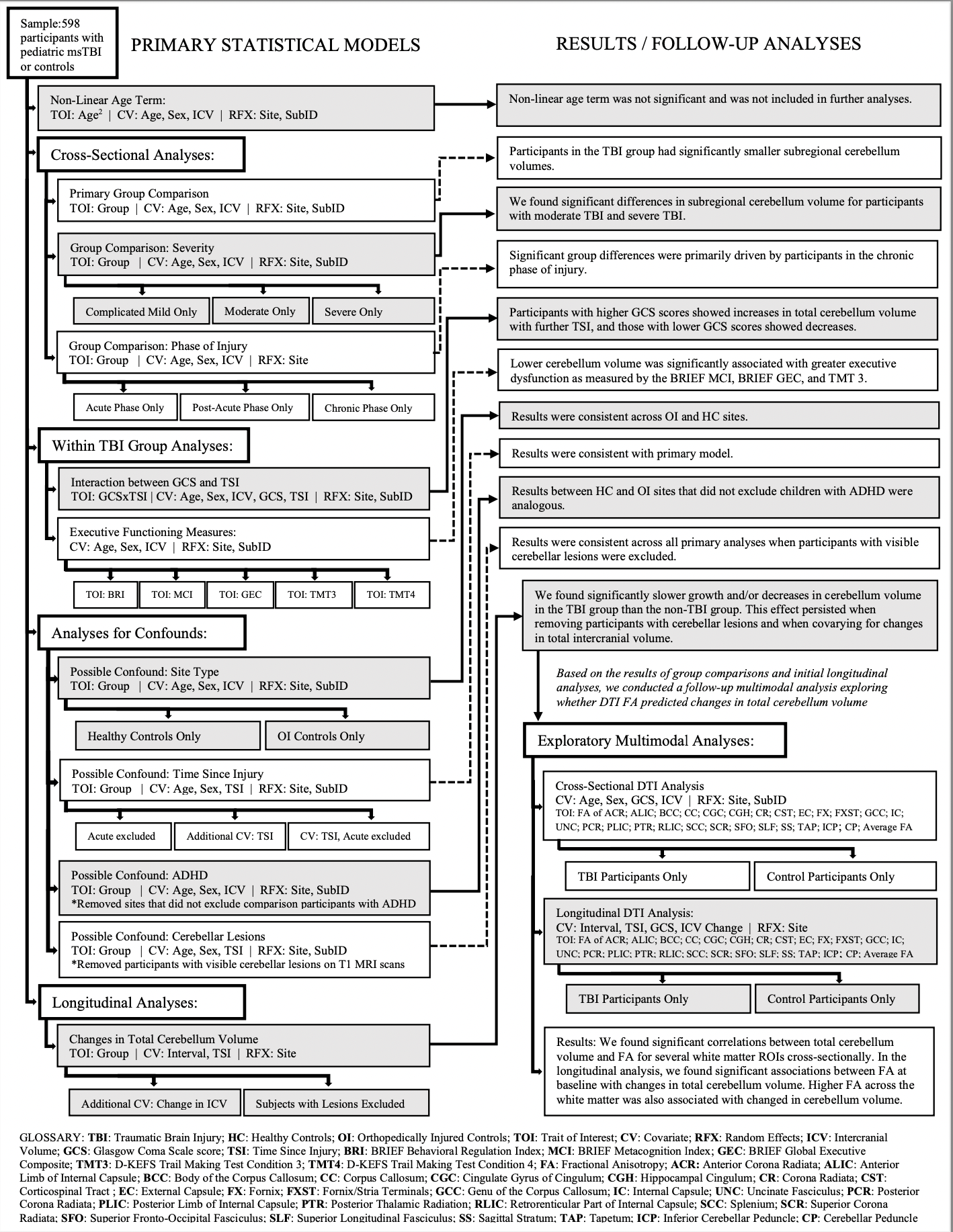

| **Supplementary Table 5**: Group comparisons in different phases of Injury | | | | | | | |  |  |  |  |  |
| --- | --- | --- | --- | --- | --- | --- | --- | --- | --- | --- | --- | --- |
|  |  | Acute | |  |  | Post-Acute | | |  | Chronic | |  |
| Region | Subregion | *p*-value | Cohen’s D | CI |  | *p*-value | Cohen’s D | CI |  | *p*-value | Cohen’s D | CI |
| Total Volume |  | 0.12 | -0.28 | [-0.63,0.07] |  | 0.055 | -0.25 | [-0.50,0.00] |  | **6.5E-08** | **-0.55** | **[-0.75,-0.35]** |
| Corpus Medullare |  | 3.0E-02* | -0.39* | [-0.75,-0.04] |  | 0.073 | -0.23 | [-0.48,0.02] |  | **3.7E-07** | **-0.53** | **[-0.73,-0.32]** |
|  | Lobule I.III | 0.62 | 0.09 | [-0.26,0.44] |  | 0.57 | 0.07 | [-0.18,0.33] |  | 0.024* | -0.23* | [-0.43,-0.03] |
| Anterior Lobe | Lobule IV | 0.49 | 0.12 | [-0.23,0.47] |  | 0.84 | -0.03 | [-0.28,0.23] |  | 0.49 | 0.07 | [-0.13,0.27] |
|  | Lobule V | 0.98 | -0.01 | [-0.36,0.35] |  | 0.84 | -0.03 | [-0.28,0.23] |  | **3.5E-03** | **-0.30** | **[-0.50,-0.10]** |
|  | Lobule VI | 0.16 | -0.25 | [-0.60,0.10] |  | 0.64 | -0.07 | [-0.35,0.22] |  | 0.025* | -0.24* | [-0.45,-0.03] |
|  | Crus I | 0.49 | 0.13 | [-0.24,0.49] |  | 0.042* | -0.33 | [-0.64,-0.01] |  | 0.078 | -0.21 | [-0.44,0.02] |
|  | Crus II | 0.10 | -0.31 | [-0.68,0.06] |  | 0.096 | -0.24 | [-0.51,0.04] |  | **2.7E-04** | **-0.40** | **[-0.62,-0.19]** |
| Posterior Lobe | Lobule VIIB | 0.13 | -0.28 | [-0.64,0.08] |  | 0.61 | -0.07 | [-0.33,0.20] |  | 0.017* | -0.26* | [-0.46,-0.05] |
|  | Lobule VIIIA | 0.75 | -0.06 | [-0.42,0.30] |  | 0.54 | 0.09 | [-0.19,0.36] |  | 0.024* | -0.26* | [-0.48,-0.03] |
|  | Lobule VIIIB | 0.54 | -0.12 | [-0.48,0.25] |  | 1.9E-02* | -0.35* | [-0.65,-0.06] |  | **1.5E-06** | **-0.55** | **[-0.77,-0.32]** |
|  | Lobule IX | 0.82 | 0.04 | [-0.32,0.40] |  | 0.11 | -0.23 | [-0.50,0.05] |  | 0.052 | -0.21 | [-0.41,0.00] |
| Flocculonodular Lobe | Lobule X | 0.65 | 0.08 | [-0.27,0.43] |  | 0.87 | 0.02 | [-0.23,0.27] |  | 0.62 | 0.05 | [-0.15,0.25] |
|  | Vermis X | 0.94 | -0.01 | [-0.37,0.34] |  | 0.082 | -0.23 | [-0.48,0.03] |  | 0.016* | -0.24* | [-0.44,-0.05] |
|  | Vermis VI | 0.26 | -0.21 | [-0.57,0.15] |  | 0.47 | 0.09 | [-0.16,0.35] |  | 0.28 | 0.11 | [-0.09,0.31] |
| Vermis | Vermis VII | **2.3E-04** | **-0.69** | **[-1.05,-0.32]** |  | 0.14 | -0.19 | [-0.45,0.06] |  | **1.4E-03** | **-0.32** | **[-0.52,-0.12]** |
|  | Vermis VIII | 0.80 | 0.04 | [-0.31,0.40] |  | 0.060 | -0.24 | [-0.49,0.01] |  | 0.23 | -0.12 | [-0.32,0.08] |
|  | Vermis IX | 0.51 | -0.12 | [-0.48,0.24] |  | 0.061 | -0.24 | [-0.50,0.01] |  | **1.1E-03** | **-0.24** | **[-0.50,0.01]** |
| The p-values, Cohen d-values, and the 95% confidence intervals for d-values are shown for results sorted by phase of injury. Acute<6 weeks post-injury, Post-acute=6 weeks to 6 months since injury, Chronic=>6 months post-injury..  Bolded values are significant (based on Li and Ji adjusted Bonferroni correction).  * Results are significant at uncorrected p<0.05 | | | | | | | | | | | | |

| **Supplementary Table 6**: Group comparisons with different TBI severity groups | | | | | | | |  |  |  |  |  |
| --- | --- | --- | --- | --- | --- | --- | --- | --- | --- | --- | --- | --- |
|  |  | Complicated Mild | |  |  | Moderate |  |  |  | Severe | |  |
| Region | Subregion | *p*-value | Cohen’s D | CI |  | *p*-value | Cohen’s D | CI |  | *p*-value | Cohen’s D | CI |
| Total Volume |  | 0.48 | -0.07 | [-0.26,0.12] |  | 9.5E-03* | -0.27* | [-0.47,-0.07] |  | **2.9E-04** | **-0.33** | **[-0.51,-0.15]** |
| Corpus Medullare |  | 0.092 | -0.17 | [-0.36,0.03] |  | **3.2E-03** | **-0.30** | **[-0.50,-0.10]** |  | **5.8E-06** | **-0.42** | **[-0.60,-0.24]** |
|  | Lobule I.III | 0.91 | -0.01 | [-0.20,0.18] |  | 0.30 | -0.11 | [-0.31,0.09] |  | 0.78 | -0.03 | [-0.20,0.15] |
| Anterior Lobe | Lobule IV | 0.86 | 0.02 | [-0.18,0.21] |  | 0.73 | 0.04 | [-0.17,0.24] |  | 0.80 | -0.02 | [-0.20,0.16] |
|  | Lobule V | 0.91 | 0.01 | [-0.18,0.20] |  | 0.083 | -0.18 | [-0.38,0.02] |  | **4.2E-03** | **-0.26** | **[-0.44,-0.08]** |
|  | Crus I | 0.93 | 9.4E-03 | [-0.22,0.23] |  | 0.83 | -0.03 | [-0.26,0.21] |  | **0.0030** | **-0.33** | **[-0.54,-0.11]** |
|  | Crus II | 0.13 | -0.16 | [-0.37,0.05] |  | 0.013* | -0.28* | [-0.49,-0.06] |  | 5.5E-03* | -0.28* | [-0.47,-0.08] |
| Posterior Lobe | Lobule VI | 0.42 | 0.09 | [-0.12,0.29] |  | 0.57 | -0.06 | [-0.28,0.15] |  | 6.2E-03* | -0.27* | [-0.46,-0.08] |
|  | Lobule VII | 0.19 | -0.13 | [-0.33,0.07] |  | **3.7E-04** | **-0.38** | **[-0.59,-0.17]** |  | 0.016* | -0.23* | [-0.42,-0.04] |
|  | Lobule VIIIA | 0.77 | -0.03 | [-0.24,0.18] |  | 0.30 | -0.12 | [-0.34,0.10] |  | 0.97 | 0.00 | [-0.19,0.20] |
|  | Lobule VIIIB | 0.021* | -0.26* | [-0.47,-0.04] |  | 0.32 | -0.12 | [-0.34,0.11] |  | **2.6E-04** | **-0.38** | **[-0.58,-0.18]** |
|  | Lobule IX | 0.12 | -0.16 | [-0.37,0.04] |  | 0.56 | -0.06 | [-0.28,0.15] |  | 0.010* | -0.25* | [-0.44,-0.06] |
| Flocculonodular Lobe | Lobule X | 0.44 | -0.08 | [-0.27,0.12] |  | 0.64 | -0.05 | [-0.25,0.15] |  | 0.52 | -0.06 | [-0.24,0.12] |
|  | Vermis X | 0.22 | -0.12 | [-0.31,0.07] |  | 0.24 | -0.12 | [-0.32,0.08] |  | 0.014* | -0.23* | [-0.41,-0.05] |
|  | Vermis VI | 0.36 | 0.09 | [-0.10,0.29] |  | 0.26 | 0.12 | [-0.09,0.32] |  | 0.54 | 0.06 | [-0.12,0.24] |
| Vermis | Vermis VII | 0.11 | -0.16 | [-0.35,0.04] |  | 0.043* | -0.21* | [-0.41,-0.01] |  | 0.063 | -0.17 | [-0.35,0.01] |
|  | Vermis VIII | 0.19 | -0.13 | [-0.32,0.06] |  | 0.44 | -0.08 | [-0.28,0.12] |  | 0.085 | -0.16 | [-0.34,0.02] |
|  | Vermis IX | 0.76 | -0.03 | [-0.22,0.16] |  | 0.18 | -0.14 | [-0.34,0.06] |  | 0.034* | -0.19* | [-0.37,-0.01] |
| The p-values, Cohen d-values, and the 95% confidence intervals for d-values are shown for results sorted by GCS severity. Complicated Mild=13-15, Moderate=9=12, Severe=3-8  Bolded values are significant (based on Li and Ji adjusted Bonferroni correction).  * Results are significant at uncorrected p<0.05 | | | | | | | | | | | | |

**Supplementary Figure 2.** Association between age at injury and percent change in total cerebellum volume for the subset of TBI participants with longitudinal data available.

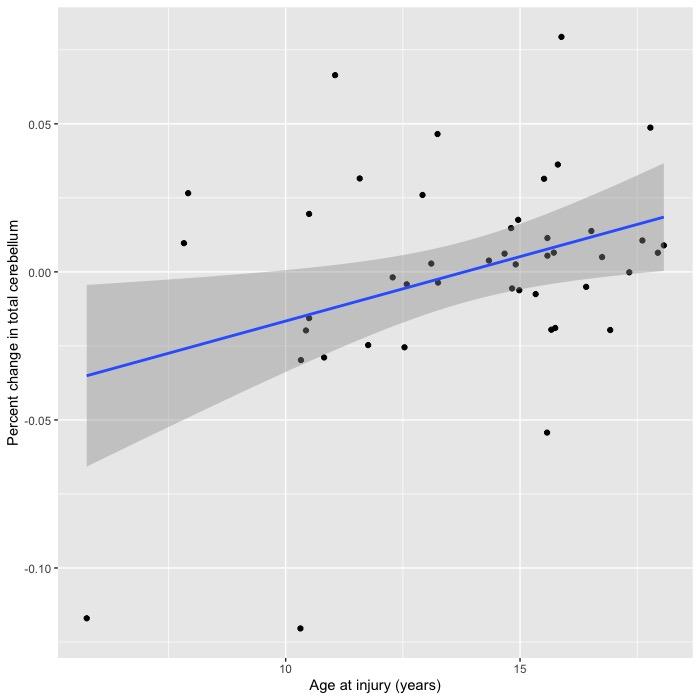

| **Supplementary Table 7**: **Supplemental Group Comparisons.** Results are shown for the main group comparison 1) excluding participants within 7 weeks of injury, 2) excluding participants within 7 weeks of injury and covarying for time since injury (TSI), and 3) covarying for TSI. | | | | | | | | | | | | |
| --- | --- | --- | --- | --- | --- | --- | --- | --- | --- | --- | --- | --- |
|  |  | Post-Acute / Chronic Only | | |  | Post Acute / Chronic Only + covaried for TSI | | |  | All Phases + covaried for TSI | | |
| Region | Subregion | *p*-value | Cohen’s D | CI |  | *p*-value | Cohen’s D | CI |  | *p*-value | Cohen’s D | CI |
| Total Volume |  | **9.4E-08** | **-0.46** | **[-0.62,-0.29]** |  | **4.4E-05** | **-0.37** | **[-0.55,-0.19]** |  | **7.5E-06** | **-0.37** | **[-0.53,-0.21]** |
| Corpus Medullare |  | **9.2E-06** | **-0.38** | **[-0.55,-0.21]** |  | **1.2E-03** | **-0.30** | **[-0.47,-0.12]** |  | **6.7E-04** | **-0.28** | **[-0.44,-0.12]** |
|  | Lobule I.III | 0.11 | -0.14 | [-0.30,0.03] |  | 0.47 | -0.07 | [-0.24,0.11] |  | 0.74 | -0.03 | [-0.19,0.13] |
| Anterior Lobe | Lobule IV | 0.73 | 0.03 | [-0.14,0.20] |  | 0.75 | 0.03 | [-0.15,0.21] |  | 1.0 | 0.00 | [-0.16,0.16] |
|  | Lobule V | 0.012* | -0.21* | [-0.38,-0.05] |  | 0.15 | -0.13 | [-0.31,0.05] |  | 0.14 | -0.12 | [-0.28,0.04] |
|  | Crus I | 0.0046* | -0.3 | [-0.50,-0.09] |  | 0.0066* | -0.3 | [-0.51,-0.08] |  | 0.0090* | -0.25 | [-0.44,-0.06] |
|  | Crus II | **1.3E-04** | **-0.36** | **[-0.54,-0.18]** |  | **4.9E-03** | **-0.28** | **[-0.47,-0.08]** |  | 5.0E-03* | -0.25* | [-0.42,-0.08] |
| Posterior Lobe | Lobule VI | 0.050 | -0.18 | [-0.36,0.00] |  | 0.35 | -0.09 | [-0.29,0.10] |  | 0.032* | -0.19* | [-0.36,-0.02] |
|  | Lobule VII | **1.8E-03** | **-0.28** | **[-0.46,-0.10]** |  | **3.3E-03** | **-0.28** | **[-0.47,-0.09]** |  | **3.0E-03** | **-0.25** | **[-0.42,-0.09]** |
|  | Lobule VIIIA | 0.60 | -0.05 | [-0.24,0.14] |  | 0.57 | -0.06 | [-0.28,0.15] |  | 0.64 | -0.04 | [-0.22,0.14] |
|  | Lobule VIIIB | **1.2E-05** | **-0.43** | **[-0.62,-0.24]** |  | **2.5E-04** | **-0.38** | **[-0.59,-0.18]** |  | **1.4E-04** | **-0.35** | **[-0.54,-0.17]** |
|  | Lobule IX | 0.058 | -0.17 | [-0.35,0.01] |  | 0.037* | -0.20* | [-0.39,-0.01] |  | 0.064 | -0.16 | [-0.33,0.01] |
| Flocculonodular Lobe | Lobule X | 0.99 | 0.00 | [-0.17,0.17] |  | 0.69 | -0.04 | [-0.21,0.14] |  | 0.89 | -0.01 | [-0.17,0.15] |
|  | Vermis X | **1.5E-03** | **-0.27** | **[-0.44,-0.10]** |  | **4.1E-04** | **-0.32** | **[-0.50,-0.14]** |  | **1.0E-03** | **-0.27** | **[-0.43,-0.11]** |
|  | Vermis VI | 0.11 | 0.13 | [-0.03,0.30] |  | 0.14 | 0.13 | [-0.04,0.31] |  | 0.58 | 0.05 | [-0.12,0.21] |
| Vermis | Vermis VII | 0.083 | -0.15 | [-0.31,0.02] |  | 0.092 | -0.15 | [-0.33,0.02] |  | **4.5E-03** | **-0.23** | **[-0.40,-0.07]** |
|  | Vermis VIII | 0.10 | -0.14 | [-0.30,0.03] |  | 0.017* | -0.22* | [-0.39,-0.04] |  | 0.018* | -0.19* | [-0.35,-0.03] |
|  | Vermis IX | 0.013* | -0.21* | [-0.38,-0.05] |  | 0.057 | -0.17 | [-0.35,0.01] |  | 0.015* | -0.20* | [-0.36,-0.04] |
| The p-values, Cohen d-values, and the 95% confidence intervals for d-values are shown for supplemental group comparison models.  Bolded values are significant (based on Li and Ji adjusted Bonferroni correction).  * Results are significant at uncorrected p<0.05 | | | | | | | | | | | | |

| **Supplementary Table 8**: Group Comparisons with different control groups | | | | | | | |  |  |  |  |  |  |  |
| --- | --- | --- | --- | --- | --- | --- | --- | --- | --- | --- | --- | --- | --- | --- |
|  |  | Healthy Controls | |  |  | OI Comparisons | | | Healthy Controls (no ADHD) | | | OI Comparisons (no ADHD) | | |
| Region | Subregion | *p*-value | Cohen’s D | CI |  | *p*-value | Cohen’s D | CI | *p*-value | Cohen’s D | CI | *p*-value | Cohen’s D | CI |
| Total Volume |  | **4.2E-08** | **-0.42** | **[-0.60,-0.24]** |  | 0.018* | -0.13* | [-0.38,0.12] | **5.2E-07** | **-0.64** | **[-0.88,-0.39]** | 0.014 | -0.38 | [-0.69,-0.08] |
| Corpus Medullare |  | **6.6E-09** | **-0.33** | **[-0.51,-0.15]** |  | 0.036* | -0.13* | [-0.38,0.11] | **1.5E-07** | **-0.67** | **[-0.92,-0.42]** | 0.017 | -0.38 | [-0.68,-0.07] |
|  | Lobule I.III | 0.27 | -0.03 | [-0.20,0.15] |  | 0.81 | -0.05 | [-0.36,0.26] | 0.44 | -0.10 | [-0.34,0.15] | 0.43 | -0.12 | [-0.43,0.18] |
| Anterior Lobe | Lobule IV | 0.59 | -0.02 | [-0.20,0.16] |  | 0.34 | -0.11 | [-0.39,0.16] | 0.48 | -0.09 | [-0.33,0.16] | 0.22 | 0.20 | [-0.12,0.51] |
|  | Lobule V | 0.041* | -0.26* | [-0.44,-0.08] |  | 0.20 | -0.07 | [-0.32,0.17] | 0.037* | -0.26 | [-0.50,-0.02] | 0.27 | -0.17 | [-0.48,0.14] |
|  | Crus I | 0.0076* | -0.28 | [-0.49,-0.08] |  | 0.75 | -0.05 | [-0.36,0.26] | 0.0040 | -0.45 | [-0.75,-0.14] | 0.22 | -0.29 | [-0.75,0.17] |
|  | Crus II | **2.0E-06** | **-0.28** | **[-0.47,-0.08]** |  | 0.41 | 0.12 | [-0.13,0.37] | **5.0E-06** | **-0.65** | **[-0.93,-0.37]** | 0.16 | -0.25 | [-0.59,0.09] |
| Posterior Lobe | Lobule VI | **4.4E-03** | **-0.27** | **[-0.46,-0.08]** |  | 0.95 | -0.16 | [-0.41,0.09] | 0.010* | -0.33 | [-0.58,-0.08] | 0.66 | 0.09 | [-0.31,0.49] |
|  | Lobule VII | 0.014* | -0.23* | [-0.42,-0.04] |  | 0.016* | 0.01* | [-0.28,0.30] | 0.0040* | -0.38 | [-0.64,-0.12] | 0.027* | -0.36 | [-0.67,-0.04] |
|  | Lobule VIIIA | 0.076 | 0.00 | [-0.19,0.20] |  | 0.48 | -0.33 | [-0.59,-0.06] | 0.053 | -0.26 | [-0.52,0.00] | 0.68 | 0.07 | [-0.27,0.42] |
|  | Lobule VIIIB | **8.8E-05** | **-0.38** | **[-0.58,-0.18]** |  | 0.026* | 0.11* | [-0.19,0.41] | **8.8E-05** | **-0.53** | **[-0.79,-0.26]** | 0.16 | -0.29 | [-0.70,0.11] |
|  | Lobule IX | 0.31 | -0.25 | [-0.44,-0.06] |  | 0.045* | -0.35* | [-0.66,-0.04] | 0.38 | -0.11 | [-0.36,0.14] | 0.042* | -0.40 | [-0.78,-0.01] |
| Flocculonodular Lobe | Lobule X | 0.50 | -0.06 | [-0.24,0.12] |  | 0.56 | -0.29 | [-0.57,-0.01] | 0.20 | 0.16 | [-0.08,0.40] | 0.62 | -0.08 | [-0.38,0.23] |
|  | Vermis X | 0.057 | -0.23 | [-0.41,-0.05] |  | 0.10 | -0.26 | [-0.51,-0.02] | 0.037* | -0.26 | [-0.50,-0.02] | 0.28 | -0.17 | [-0.48,0.14] |
|  | Vermis VI | 0.62 | 0.06 | [-0.12,0.24] |  | 0.14 | -0.30 | [-0.54,-0.05] | 0.95 | -0.01 | [-0.25,0.23] | 0.12 | 0.25 | [-0.06,0.56] |
| Vermis | Vermis VII | 0.032* | -0.17* | [-0.35,0.01] |  | 0.088 | -0.21 | [-0.46,0.04] | 0.49 | -0.09 | [-0.33,0.16] | 0.18 | -0.21 | [-0.52,0.10] |
|  | Vermis VIII | 0.071 | -0.16 | [-0.34,0.02] |  | 0.29 | 0.19 | [-0.06,0.44] | 0.026* | -0.27 | [-0.52,-0.03] | 0.18 | -0.21 | [-0.52,0.10] |
|  | Vermis IX | **2.2E-03** | **-0.19** | **[-0.37,-0.01]** |  | 0.31 | -0.22 | [-0.47,0.03] | 0.015* | -0.30 | [-0.55,-0.06] | 0.89 | 0.02 | [-0.29,0.33] |
| The p-values, Cohen d-values, and the 95% confidence intervals for d-values are shown for results sorted by site type.  Bolded values are significant (based on Li and Ji adjusted Bonferroni correction).  * Results are significant at uncorrected p<0.05 | | | | | | | | |  |  |  |  |  |  |

**Supplementary Table 9.** Cross-sectional and longitudinal associations between FA and total cerebellum volume in the TBI group. Longitudinal associations are between FA at first scan and changes in total cerebellum volume. FA=fractional anisotropy, ACR=anterior corona radiata, ALIC=anterior limb of internal capsule, BCC=body of corpus callosum, CC=corpus callosum, CGC=cingulum, CGH=hippocampal cingulum, CP=cerebellar peduncle, CR=corona radiata, CST=corticospinal tract, EC=external capsule, FX=fornix, FXST=fornix stria terminalis, GCC=genu of corpus callosum, IC=internal capsule, ICP=inferior cerebellar peduncle, PCR=posterior corona radiata, PLIC=posterior limb of internal capsule, PTR=posterior thalamic radiation, RLIC=retrolenticular limb of internal capsule, SCC=splenium of corpus callosum, SCP=superior cerebellar peduncle, SCR=superior corona radiata, SFO=superior fronto-occipital fasciculus, SLF=superior longitudinal fasciculus, SS=sagittal stratum, TAP=tapetum, UNC=uncinate. Significant associations are **bolded**.

|  | **Cross-sectional, covarying for GCS (N=252)** | | **Longitudinal, covarying for interval, TSI, and GCS (N=32)** | |
| --- | --- | --- | --- | --- |
| **ROI** | ***t-value*** | ***p-value*** | ***t-value*** | ***p-value*** |
| AverageFA | **4.1** | **1.7E-04** | **3.2** | **4.9E-03** |
| ACR | 1.6 | 0.12 | **2.3** | **0.030** |
| ALIC | **1.9** | **0.064** | 2.0 | 0.061 |
| BCC | **3.4** | **1.5E-03** | **3.1** | **5.8E-03** |
| CC | **3.6** | **6.9E-04** | **2.9** | **8.3E-03** |
| CGC | **3.6** | **6.4E-04** | **3.1** | **5.4E-03** |
| CGH | 1.9 | 0.068 | **2.3** | **0.034** |
| CP | **2.7** | **0.010** | 0.3 | 0.76 |
| CR | **2.3** | **0.027** | **2.3** | **0.031** |
| CST | **2.9** | **5.8E-03** | 0.9 | 0.36 |
| EC | **2.8** | **8.1E-03** | 1.6 | 0.13 |
| FX | **3.5** | **1.0E-03** | **3.8** | **1.0E-03** |
| FXST | **2.9** | **5.2E-03** | 1.2 | 0.23 |
| GCC | 1.4 | 0.16 | 1.1 | 0.29 |
| IC | **2.2** | **0.029** | 1.8 | 0.096 |
| ICP | **2.6** | **0.013** | **2.5** | **0.021** |
| PCR | 1.9 | 0.057 | **2.3** | **0.030** |
| PLIC | 1.4 | 0.16 | 1.0 | 0.32 |
| PTR | **2.4** | **0.019** | **2.4** | **0.025** |
| RLIC | **2.8** | **6.5E-03** | **3.2** | **4.5E-03** |
| SCC | **3.8** | **4.2E-04** | **2.3** | **0.034** |
| SCP | 1.8 | 0.083 | 0.7 | 0.48 |
| SCR | **2.5** | **0.016** | 1.9 | 0.077 |
| SFO | 1.2 | 0.24 | 2.0 | 0.054 |
| SLF | **3.8** | **3.7E-04** | **2.9** | **9.2E-03** |
| SS | **2.5** | **0.0170** | **2.4** | **0.026** |
| TAP | **3.4** | **1.5E-03** | **3.3** | **3.6E-03** |
| UNC | **2.8** | **6.6E-03** | 0.0 | 0.98 |

**Supplementary Figure 3.** White matter regions associated with longitudinal changes in total cerebellum volume in the TBI group. Color corresponds to the Pearson’s *r* as depicted in the color bar.
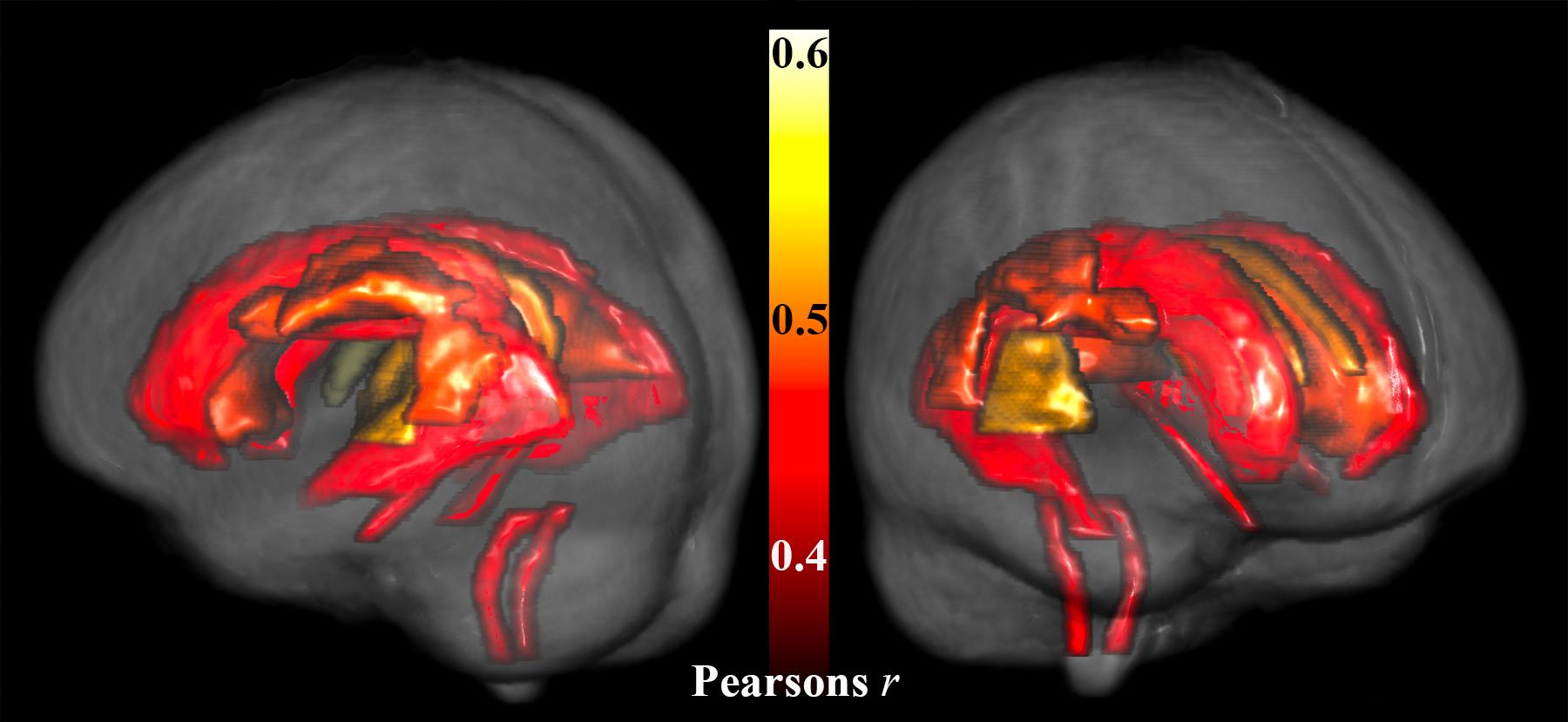

| **Supplementary Table 10**: Interactions | | | | | | | | | |  |  |  |  |  |  |
| --- | --- | --- | --- | --- | --- | --- | --- | --- | --- | --- | --- | --- | --- | --- | --- |
|  |  | Group x Age | |  | Group x Sex | |  | AgeatInj x TSI | |  | AgeatInj x GCS | |  | TSI x GCS | |
| Region | Subregion | *p*-value | *b-*value |  | *p*-value | *b-*value |  | *p*-value | *b-*value |  | *p*-value | *b-*value |  | *p*-value | *b-*value |
| Total Volume |  | 0.69 | 136.03 |  | 0.36 | -1952.13 |  | 0.34 | -0.050 |  | 0.074 | -24.53 |  | **1.8E-04** | **6.33** |
| Corpus Medullare |  | 0.37 | -72.12 |  | 0.33 | -496.09 |  | 0.76 | 0.060 |  | 0.79 | 2.20 |  | 0.049* | 0.76 |
|  | Lobule I.III | 0.30 | 6.33 |  | 0.058 | -71.05 |  | 0.71 | 0.020 |  | 0.76 | -0.11 |  | 0.52 | -0.02 |
| Anterior Lobe | Lobule IV | 0.85 | -2.82 |  | 0.90 | 12.30 |  | 0.48 | -0.010 |  | 0.83 | 0.31 |  | 0.13 | 0.11 |
|  | Lobule V | 0.13 | 19.18 |  | 0.55 | 46.60 |  | 0.37 | -0.040 |  | 0.86 | -0.47 |  | 0.22 | 0.09 |
|  | Crus I | 0.39 | -56.90 |  | 0.60 | -214.70 |  | 0.81 | -0.046 |  | 0.014* | -24.50 |  | **0.0025** | **1.0** |
|  | Crus II | 0.94 | -3.13 |  | 0.46 | 189.56 |  | 0.65 | 0.22 |  | 0.74 | -5.28 |  | 0.93 | 0.02 |
| Posterior Lobe | Lobule VI | 0.45 | 27.70 |  | 0.40 | -187.92 |  | 0.072 | 0.15 |  | 0.30 | -6.54 |  | 0.014* | 0.42 |
|  | Lobule VII | 0.14 | 48.69 |  | 0.83 | 43.63 |  | 0.25 | -0.12 |  | 0.25 | 0.39 |  | 0.010* | 0.52 |
|  | Lobule VIIIA | 0.20 | -43.13 |  | 0.75 | -65.60 |  | 0.31 | -0.070 |  | 0.94 | 1.40 |  | 0.12 | 0.34 |
|  | Lobule VIIIB | 0.94 | -1.42 |  | 0.45 | -89.77 |  | 0.36 | 0.050 |  | 0.65 | -1.37 |  | 0.039* | 0.25 |
|  | Lobule IX | 0.81 | 4.76 |  | 0.45 | -91.81 |  | 0.43 | 0.080 |  | 0.65 | -3.37 |  | 0.053 | 0.17 |
| Flocculonodular Lobe | Lobule X | 0.13 | 4.36 |  | 0.14 | -26.03 |  | 0.040* | 1.11 |  | 0.80 | -100.16 |  | 0.41 | 0.01 |
|  | Vermis X | 0.23 | 2.18 |  | 0.61 | -5.85 |  | 0.57 | 0.00 |  | 0.97 | 0.010 |  | 0.92 | 0.00 |
|  | Vermis VI | 0.13 | 10.86 |  | 0.098 | -72.99 |  | 0.47 | 0.020 |  | 0.91 | 0.13 |  | 0.32 | 0.04 |
| Vermis | Vermis VII | 0.12 | 7.87 |  | 0.66 | 13.96 |  | 0.95 | 0.00 |  | 0.68 | 0.30 |  | 0.20 | 0.03 |
|  | Vermis VIII | 0.81 | -2.16 |  | 0.064 | -104.43 |  | 0.78 | -0.010 |  | 0.026* | -3.25 |  | 0.27 | 0.04 |
|  | Vermis IX | 0.016* | 10.60 |  | 0.53 | 17.17 |  | 0.11 | 0.030 |  | 0.14 | -1.02 |  | 0.37 | 0.02 |
| The p-values and beta-values are shown for interaction models.  Bolded values are significant (based on Li and Ji adjusted Bonferroni correction).  * Results are significant at uncorrected p<0.05 | | | | | | | | | |  |  |  |  |  |  |

**Supplementary Figure 4. Interaction between time since injury and injury severity.** The interaction between time since injury in weeks and Glasgow Coma Scale is shown for total cerebellum volume (normalized residuals accounting for age, sex, ICV, and random effects of site and subject). TSI is truncated at 4 years. Light blue dots and dotted line are for participants with GCS in the bottom tertile (most severe), dark blue dots and solid line are for participants with GCS in the top tertile (least severe), with the middle blue dots and dashed line for participants in the middle tertile.
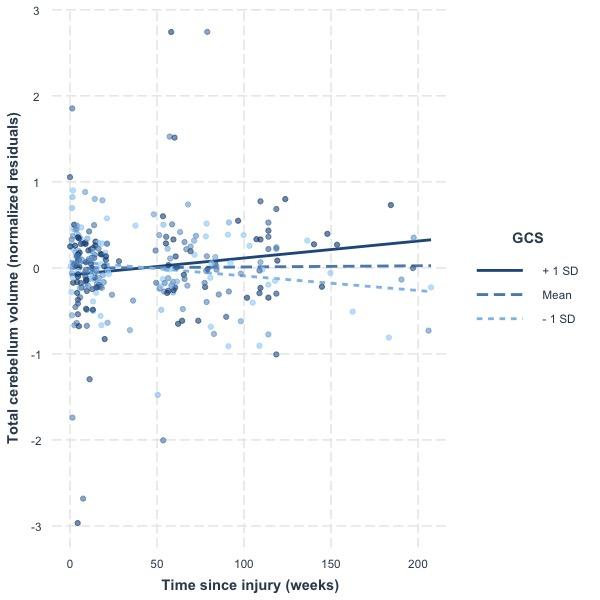

| **Supplementary Table 11**: D-KEFS Trail Making Test Scores | | | | | | | |  |  |
| --- | --- | --- | --- | --- | --- | --- | --- | --- | --- |
|  |  |  | TMT 3 | | |  | TMT 4 | | |
| Region | Subregion |  | *p*-value | Adj-*p* | *b*-value |  | *p*-value | Adj-*p* | *b*-value |
| Total Volume |  |  | 0.0080* | 0.085 | 3.16 |  | 0.017* | 0.17 | 2.81 |
| Corpus Medullare |  |  | 0.069 | 0.54 | 2.01 |  | 0.038* | 0.35 | 2.39 |
|  | Lobule I-III |  | **0.0020** | 0.022 | **4.00** |  | 0.26 | 0.96 | 1.18 |
| Anterior Lobe | Lobule IV |  | 0.53 | 1.0 | 0.64 |  | 0.87 | 1.0 | -0.16 |
|  | Lobule V |  | 0.65 | 1.0 | 0.47 |  | 0.85 | 1.0 | 0.19 |
|  | Crus I |  | 0.042* | 0.38 | 78.1 |  | 0.036* | 0.33 | 135.6 |
|  | Crus II |  | 0.36 | 1.0 | 0.97 |  | 0.22 | 0.94 | 1.33 |
| Posterior Lobe | Lobule VI |  | 0.21 | 0.93 | 1.34 |  | 0.050 | 0.43 | 2.26 |
|  | Lobule VII |  | 0.28 | 0.97 | 1.13 |  | 0.42 | 1.0 | 0.83 |
|  | Lobule VIIIA |  | 0.13 | 0.79 | 1.66 |  | 0.26 | 0.96 | 1.21 |
|  | Lobule VIIIB |  | 0.046* | 0.40 | 2.24 |  | 0.10 | 0.70 | 1.78 |
|  | Lobule IX |  | 0.020* | 0.20 | 2.73 |  | 0.025* | 0.24 | 2.63 |
| Flocculonodular Lobe | Lobule X |  | 0.11 | 0.73 | 1.71 |  | 0.036* | 0.33 | 2.39 |
|  | Vermis X |  | 0.85 | 1.0 | 0.19 |  | 0.82 | 1.0 | 0.24 |
|  | Vermis VI |  | 0.72 | 1.0 | 0.37 |  | 0.63 | 1.0 | 0.50 |
| Vermis | Vermis VII |  | 0.24 | 0.95 | 1.25 |  | 0.25 | 0.96 | 1.23 |
|  | Vermis VIII |  | 0.18 | 0.89 | 1.43 |  | 0.0090* | 0.095 | 3.19 |
|  | Vermis IX |  | 0.25 | 0.96 | 1.22 |  | 0.32 | 0.99 | 1.03 |
| The p-values, adjusted p-values,and beta-values are shown for DKEFS TRT scores.  Bolded values are significant (based on Li and Ji adjusted Bonferroni correction).  * Results are significant at uncorrected p<0.05 | | | | | | | | | |
